# Probabilistic classification of late treatment failure in uncomplicated malaria

**DOI:** 10.1101/2025.01.21.25320790

**Authors:** Somya Mehra, Aimee R Taylor, Mallika Imwong, Nicholas J White, James A Watson

## Abstract

Distinguishing treatment failure (recrudescence) from reinfection in uncomplicated falciparum malaria is essential for characterising antimalarial treatment efficacy in malaria endemic areas. Classification of recrudescence versus reinfection is usually based on a comparison of parasite allelic calls derived from PCR amplification and electrophoresis of individual polymorphic markers in the acute and recurrent isolates. Match-counting methods (e.g. 3/3 or 2/3 matching alleles) have usually been applied, but these do not account for multiple comparisons per-marker when infections are polyclonal. We show that when infections are polyclonal, as is common in high transmission settings, currently used match-counting and model-based methods have unacceptably high false-discovery rates leading to overestimation of treatment failure. We develop the software *PfRecur* which provides analytical Bayesian posterior probabilities of treatment failure in recurrent falciparum malaria. We use data from a recent study in Angola to demonstrate the potential utility of our model in resolving complex polyclonal *P. falciparum* infections, thereby providing more accurate estimation of treatment failure rates.

## Introduction

*Plasmodium falciparum* is estimated to cause around 250 million symptomatic malaria cases each year, the majority of which are in young children in sub-Saharan Africa [1]. The primary therapeutic goal of antimalarial treatment for uncomplicated malaria is to cure the infection. Effective treatment of uncomplicated malaria currently relies on artemisinin-based combination therapies (ACTs). The ACTs combine a rapidly acting but rapidly eliminated artemisinin derivative with a less active and slowly eliminated partner drug (for example, the combination of artemether and lumefantrine, AL). Artemisinin resistance is now widespread in Southeast Asia [2, 3] and recently has emerged independently in East Africa [4, 5]. Resistance to artemisinin results in an increased likelihood that the ACT fails to clear the blood stage infection and recrudesces. This amplifies the selective advantage of artemisinin resistant parasites and helps select for partner drug resistance. Characterising ACT failure rates is essential to guide policies and practices. This assessment is routinely performed through therapeutic efficacy surveillance studies. These are usually single arm observational studies which enrol symptomatic uncomplicated malaria patients and follow them for one to two months after treatment. The proportion of patients with a recrudescent infection (i.e., treatment failure) is then estimated [6, 7]. Treatment failure rates should not exceed 10% [1]. In areas of high transmission where the main burden of disease is in young children and where reinfection within one month is very common, estimating the antimalarial treatment failure rate relies on distinguishing recurrent bloodstream infections resulting from incomplete clearance of the incident infection (recrudescence) from new infections (reinfection) resulting from new mosquito bites [8, 9]. This relies on molecular correction methods which compare parasite genotypes between the baseline infection with those of the recurrent infection.

Classification of a recurrent infection as either a reinfection or a recrudescence is primarily done by PCR genotyping. Length polymorphisms within genes or microsatellite markers are compared in paired acute and recurrent isolates [9]. Classification is then based on the observed alleles. Most studies have used match-counting approaches, such as those recommended by the World Health Organisation (WHO) [9]. Match-counting algorithms usually take the presence of one or more shared alleles per marker in the paired isolates as evidence of recrudescence, and can either be strict (requiring matches at all markers) or relaxed (requiring matches at a subset of markers). However, this does not account sufficiently for multiple comparisons when there are multiclonal isolates (infections caused by multiple parasite clones), the allele specific sensitivity of the genotyping method [10], the relative frequency of the different alleles, and uncertainty in the allocation of alleles across individual haploid parasite clones within each isolate [8, 11]. Concerns have been raised around the reliability of match-counting algorithms. These highlight technical constraints associated with molecular genotyping [12–14] and have been evaluated in simulation models [15, 16]. Deep sequencing of longer amplicons from SNP-rich genomic regions (AmpSeq) addresses some of these concerns, but is not yet widely available. Statistical model-based approaches can address classification uncertainty correctly and take into account background allele frequencies, adjusting for multiple comparisons. The US CDC has developed a Bayesian classifier in which allelic states across clones are explicitly estimated using a Gibbs sampler and the likelihood of recrudescence is averaged over each pairwise comparison of clones in the baseline and recurrent isolate [8]. An informal consultation convened by WHO in 2021 recommended study of “the impact of Bayesian analysis on recrudescence rates in areas of high transmission, given the trend in increased recrudescence rates using this method, to determine whether this is an artefact of the method, whether there is some reason for higher failure rates in these areas (e.g., a high MOI may be more challenging for antimalarial drugs to clear), or whether there is emergence of true antimalarial resistance that needs rigorous confirmation” [9].

In this work, we use microsatellite data provided from 70 paired *P. falciparum* recurrent infections from a study conducted in Angola in 2021 [17] to evaluate the CDC Bayesian classifier and explore the impact of model misspecification on recurrence classification. We formulate an alternative probabilistic Bayesian classifier (implemented in the R software *PfRecur*) for genotyping data from paired baseline and recurrent falciparum infections, allowing recurrent infections to be mixtures of recrudescent and newly-inoculated clones [9, 18]. We show that this formulation makes *PfRecur* robust to misspecification in the likelihood and solves combinatorial problems for multiclonal isolates, both for the pairwise comparisons between baseline and recurrent isolates [19], and the computation of sample allele frequencies [20–24]. Because the posterior is analytically tractable, *PfRecur* is fit to data analytically, precluding the need for a optimisation algorithm or numerical sampler. We compare the empirical performance of simple match-counting algorithms versus the CDC Bayesian classifier and our novel approach *PfRecur*. We show that in high transmission areas, where infections are often polyclonal, the CDC Bayesian classifier and match-counting approaches (as recommended by [16, 17, 25]) both give unacceptably high false positive rates. This can result in overestimation of treatment failure rates and thus overestimation of antimalarial drug resistance, potentially prompting expensive changes in treatment policy [26, 27].

## Results

### *PfRecur* : a probabilistic model of recrudescence

We developed a novel probabilistic model for recrudescence versus reinfection based on observed alleles from each marker in each baseline and paired recurrent infection; for simplicity we refer to this model as *PfRecur*. Rather than treating reinfection and recrudescence as mutually exclusive categories, each recurrent infection is modelled as a *mixture* of newly-inoculated (from reinfection) and recrudescent clones. This construction is motivated by a concern for robustness against model misspecification. Under the simplifying assumptions of marker-wise and clone-wise independence (within isolates and between non-recrudescent clones across isolates), we derive an analytic per-participant likelihood that averages over all allelic configurations that are compatible with the isolate MOI and the set of observed genotypes in each isolate. We accommodate undetected clones in baseline/recurrent isolates for the participant of interest only, whereby each clone is modelled to be genotyped (i.e, detected) with probability *ω* at each marker; this yields a marker-wise truncated binomial model for the number of clones that have contributed to the observed set of genotypes at each marker. Site specific allele frequencies for newly-inoculated clones are derived under a multinomial-Dirichlet model from the baseline isolates, assuming exchangeability among study patients (excluding the patient of interest). These site specific allele frequencies are also used to impute allelic states at each marker for undetected clones in the baseline isolate for each patient. Allele frequencies in the recrudescent clones are based on the paired patient baseline clones following imputation. We adjust for genotyping error in baseline isolates, relative to the recurrent isolate of interest, through a non-parametric marker-agnostic model *δ*_*𝓁*_ governing the probability that each allele called in a baseline isolate matches an allele called in the recurrent isolate; in this study, we consider a normalised geometric error model (with respect to allele repeat lengths) with error probability *ε*, adapted to length polymorphic markers. *ω, ε* are user-specified parameters in the model with default values of 0.9 and 0.05, respectively.

Under a Bayesian framework, with a symmetric beta binomial prior for the number of newly-inoculated vs recrudescent clones within a recurrent isolate, we derive an analytical posterior distribution for the number of recrudescent clones within each recurrent isolate. We consider two posterior summary metrics: the posterior probability of there being at least one recrudescent clone in the recurrent isolate (denoted M1); and the expected proportion of recrudescent clones in the recurrent isolate (M2). These metrics are identical for recurrent isolates with a single clone (i.e., MOI=1).

### Simulation study

We conducted a simulation study to evaluate the ability of our probabilistic classifier *PfRecur* to resolve mixtures of newly-inoculated and recrudescent clones. Simulated data (Appendix B) were well-specified relative to our classification model apart from three features: non-meiotic siblings (i.e. genetically related parasites derived from independent crosses of the same parental pair) could be present within isolates (thereby violating the assumption of clone-wise independence within isolates); the observed MOI of each isolate (i.e., the maximum observed cardinality across simulated markers) could be lower than the true MOI (because of undetected clones or allelic overlap between clones); and the detection of clones in baseline isolates (used to derive allele frequencies for newly-inoculated clones) was incomplete. We considered an idealised setting, where both genotyping error probabilities *ε* and the per-clone marker-wise detection probability *ω* under which data were generated were known.

Using metric M2, our model *PfRecur* was able to recapitulate the proportion of recrudescent clones within simulated recurrences, albeit with decreasing precision for recurrences with high MOIs (Figure 1A). Metric M1 (i.e., the posterior probability of at least one recrudescent clone) was sensitive to simulated recurrences with *≥* 1 recrudescent clone but was liable to call false positive recrudescences using a threshold of 0.5 (dashed blue line) at elevated MOIs (Figure 1B). Classification using a threshold of 1*/m* for recurrent isolates with elevated MOI *m* may allow recrudescences to be called with greater specificity.

**Figure 1.**
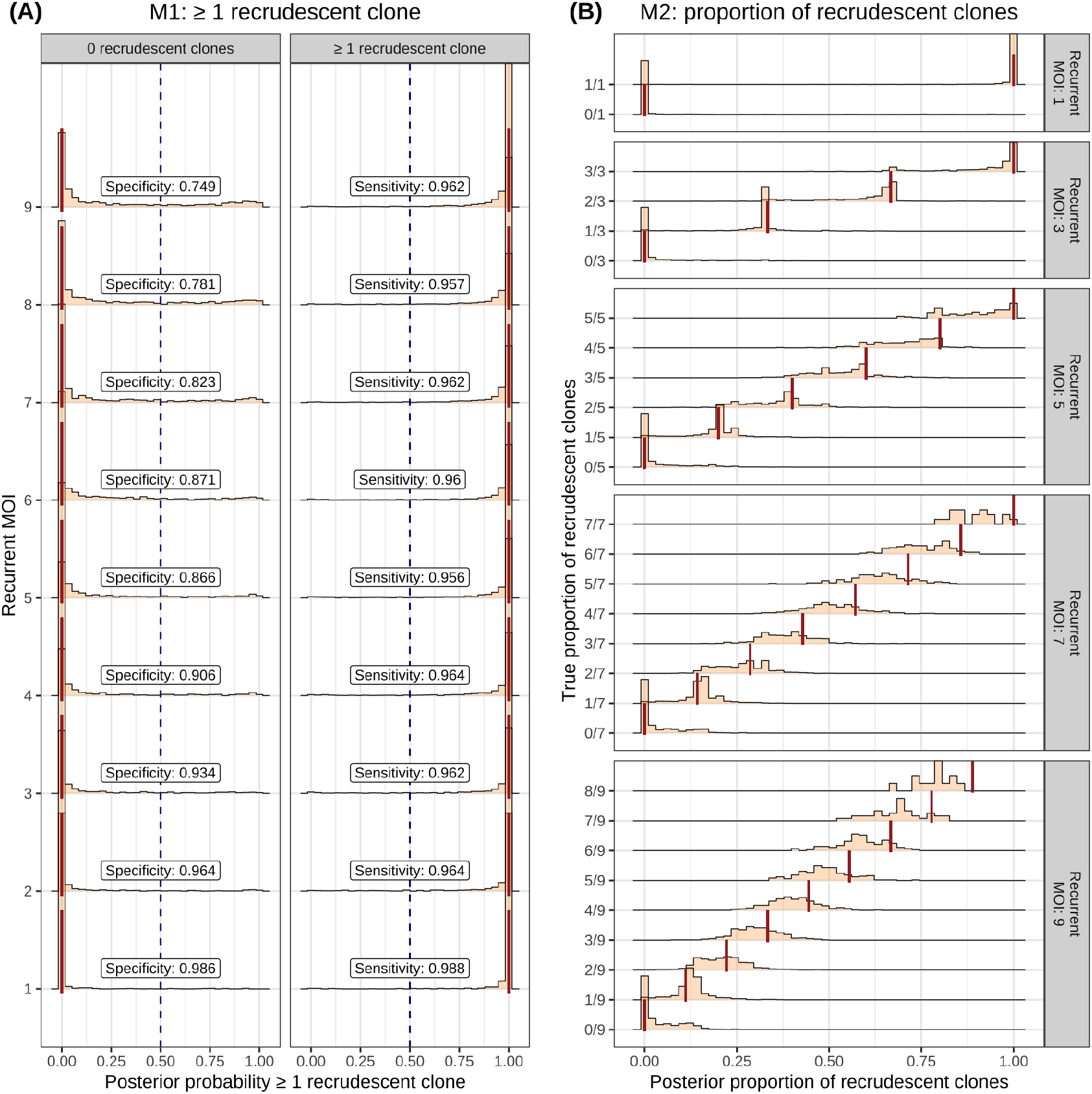
Application of our probabilistic classifier *PfRecur* to simulated mixtures of newly-inoculated and recrudescent clones. Panel A shows the posterior probability of at least one recrudescent clone (M1) (annotated with classification sensitivity and specificity using a threshold of 0.5 to call recrudescence), while panel B shows the posterior proportion of recrudescent clones (M2), aggregated over 28773 simulated recurrences. Truth values are shown with vertical lines.

### Therapeutic efficacy study in Angola

We used open access data from a therapeutic efficacy study conducted in Angola in 2021 [17] to compare *PfRecur* with match-counting algorithms and the CDC model. The Angolan study evaluated artemether-lumefantrine (AL), artesunate-amodiaquine (ASAQ), dihydroartemisinin-piperaquine (DP), and artesunate-pyronaridine (ASPY) across 3 study sites in 622 patients with uncomplicated malaria [17]. Recurrent parasitaemia was detected in 71 patients between 7 and 42 days of follow-up (Table 1). Paired baseline and recurrent *P. falciparum* isolates (70 pairs) were genotyped at 7 neutral microsatellite markers (*M2490, M313, M383, PfPK2, POLYA, TA1, TA109*). An additional 14 unpaired baseline isolates per study site were also genotyped to increase the precision of the estimation of population allele frequencies, stratified by study site. Transmission intensities varied across the three sites, with Zaire and Lunda Sul classified as high and moderate intensity transmission, respectively, and Benguela as low transmission intensity. In Zaire and Lunda Sul combined, there were 36/198 (18%) recurrent infections in the AL treated patients within 28 days of follow-up; and 19/188 (10%) recurrent infections in the artesunate-amodiaquine (ASAQ) treated patients within 28 days of follow-up. In Benguela, dihydroartemisinin-piperaquine (DP) and artesunate-pyronaridine (ASPY) were compared, with 14/100 and 6/104 recurrent infections within 42 days of follow-up, respectively. Under the CDC model, this translated into estimated day-28 PCR-corrected efficacies of 88% (95% CI 82-95) and 94.4% (95% CI 90-99) for AL; and 91% (95%CI 85-97) and 100% for ASAQ; and day-42 PCR-corrected efficacies of 99.6% (95% CI 99-100) for DP and 98.3% (95% CI 96-100) for ASPY. Of particular concern was the low efficacy estimate of AL in the Zaire site. AL is the most frequently used ACT in the world [28].

**Table 1.**
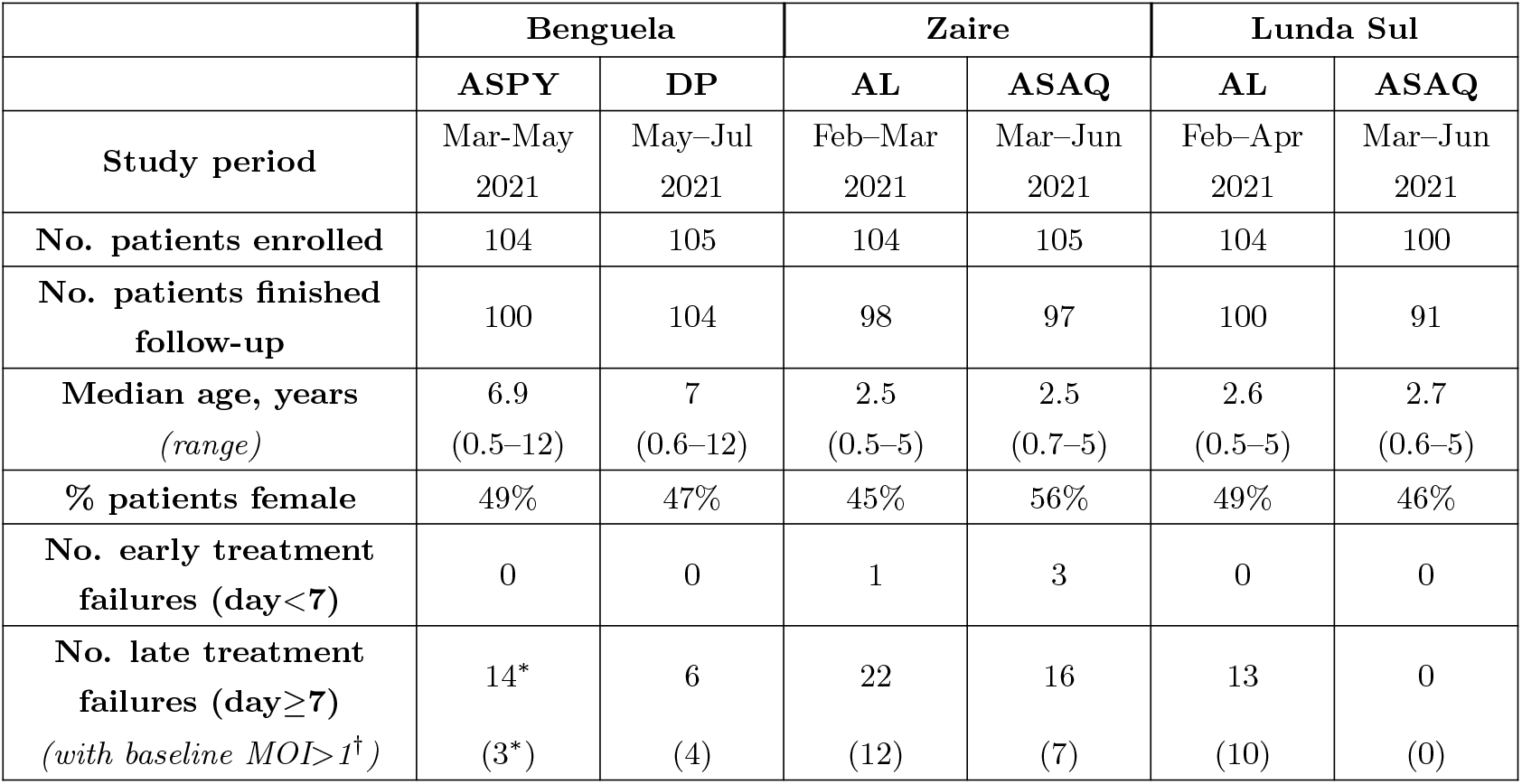
Summary of a therapeutic efficacy study conducted in Angola in 2021 by Dimbu *et al*. [17] (adapted from Tables 1 and 3 of [17]). ASPY=artesunate-pyronaridine, DP=dihydroartemisinin-piperaquine, AL=arthemether-lumenfantrine, ASAQ=artesunate-amodiaquine. ^*^Genotypes are missing for one recurrent isolate from the ASPY arm in Benguela. ^*†*^Based on the maximum observed cardinality across 7 neutral microsatellite markers for the day 0 isolate.

### Artefacts at low parasite densities

Visual inspection of the genotypic data from [17] showed that some of the markers were much more likely to be polyallelic than others. Exploratory analysis demonstrated that the parasite density (log_10_ parasitaemia) was strongly predictive of the observed MOI (maximum cardinality across the 7 markers; Figure 2A). Low densities (*<*1000 parasites per *µ*L) were associated with MOIs of 2 or more. When assessing individual markers, it was apparent that the microsatellite *TA109* is problematic particularly at very low parasite densities (Figure 2B), yielding elevated apparent MOIs for recurrent parasitaemias below 1000/*µ*L. This suggests that *TA109* multiplicity in low parasite density samples may be artefactual and result from methodological or laboratory artefacts (e.g. non-specific peaks). We conducted all analyses with and without *TA109* to test for robustness of the methodology relative to inclusion of an unreliable polymorphic marker.

**Figure 2.**
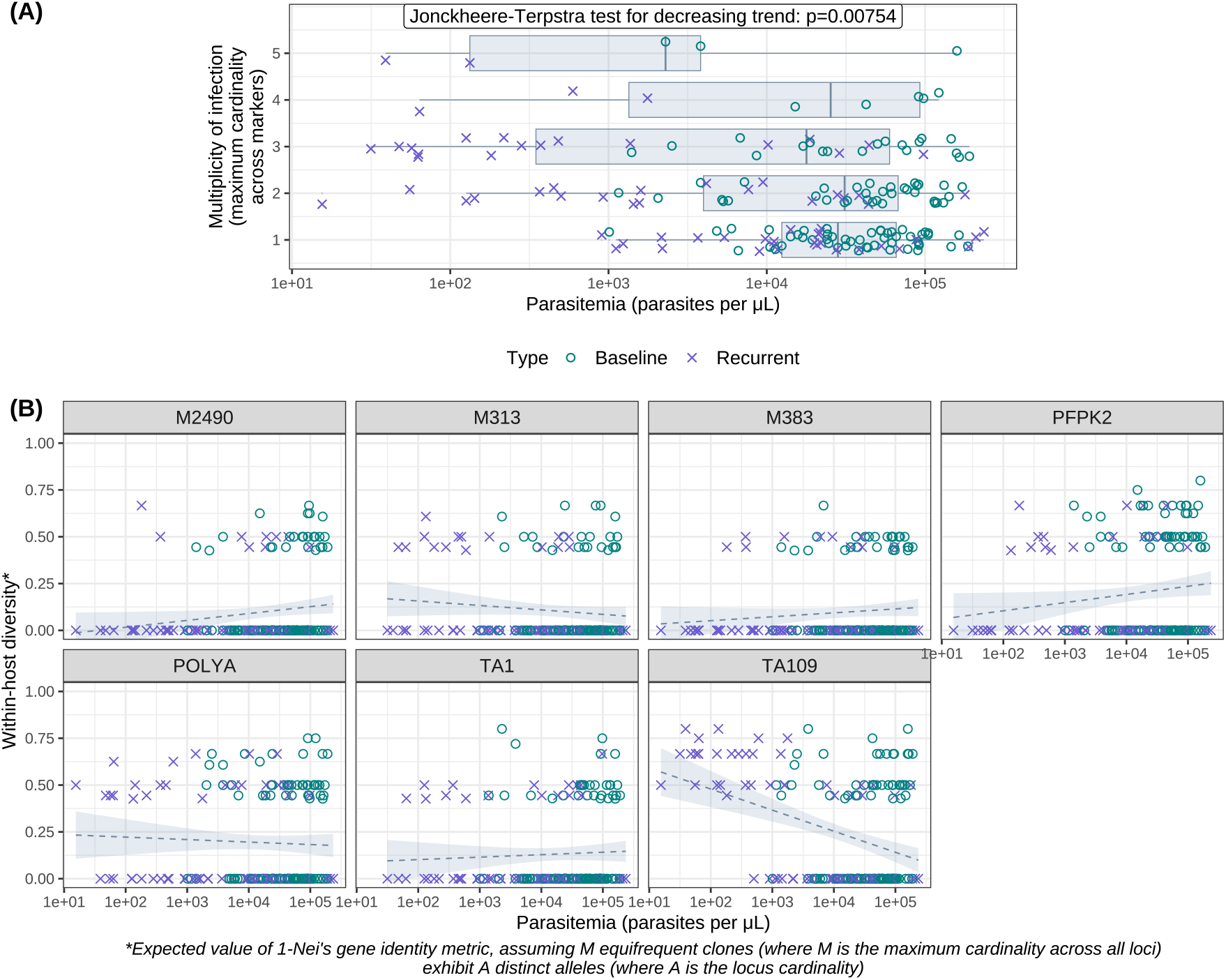
(A) Parasitemia stratified by MOI (i.e., the maximum cardinality across 7 neutral microsatel-lite markers) for 181 *P. falciparum* isolates reported by Dimbu *et al*. [17]. (B) The within-host diversity of each microsatellite marker as a function of parasitemia.

### Estimation of treatment failure for isolates with baseline MOI*>*1

We estimated the probability of recrudescence for each patient with a recurrent infection in the Angola study, using the CDC model and *PfRecur*. We used metric M1 (probability of at least one recrudescent clone) for conceptual consistency with the CDC model [8]. For paired infections with a baseline MOI of 1, the probabilistic models do not differ substantially. Both models correct for chance allelic matches, which is an advantage over a simple match-counting algorithm. However, when the MOI of the baseline exceeds 1, issues of multiple comparisons become important. Figure 3 shows the model estimates for the 36 paired infections in the Angolan study with baseline MOI*>* 1, ordered by the posterior probability of recrudescence under the CDC model. For 13/36 (one third) recurrent infections, there was a non-negligible difference in the model estimates, with the CDC model consistently estimating higher recrudescence probabilities. Discrepancies were largely apparent for recurrences with intermediate posterior probabilities under the CDC model, and many of these differences persisted after exclusion of the problematic marker *TA109*. Visual inspection of the isolates where the models differ substantially suggest that the CDC model is over-calling recrudescence (Supplementary Figure 11).

**Figure 3.**
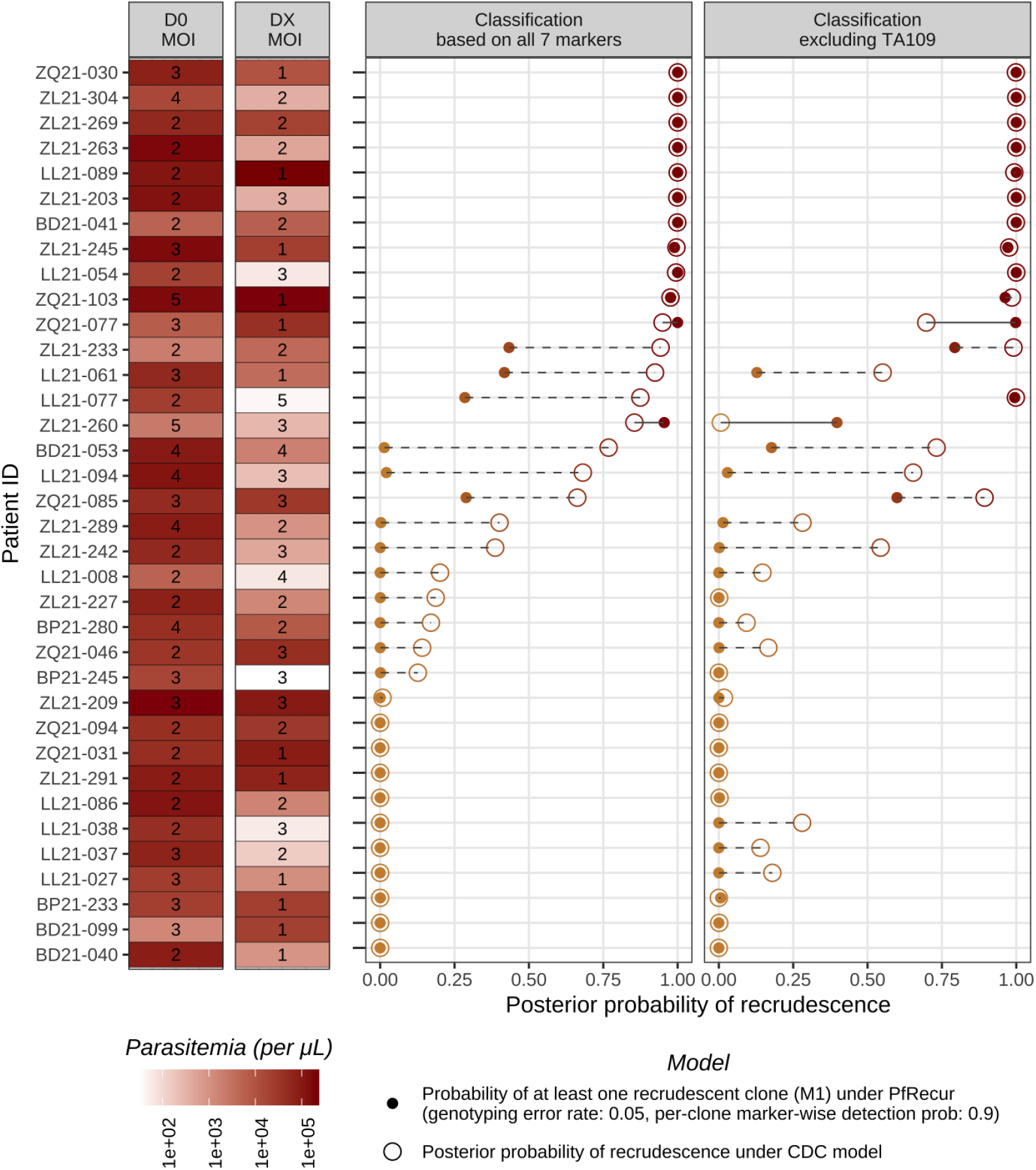
Summary of posterior estimates for *P. falciparum* recurrences reported by Dimbu *et al*. [17] with MOI*>*1 for the baseline isolate. We compare the probability of at least one recrudescent clone under *PfRecur* (with genotyping error probability *ε* = 0.05 under the normalised geometric model (29) and marker-wise per-clone detection probability *ω* = 0.9, applied to clones in the pair of baseline/recurrent isolates for the participant of interest) against the posterior probability of recrudescence based on the CDC model [8].

### Estimation of false positive rates for recrudescence classification

We used a permutation method to estimate false positive rates for recrudescence classification. We generated 500 artificial ‘not-recrudescence’ datasets by shuffling the participant identifiers of the baseline isolates in [17], stratified by study site thus preserving population structure. Figure 4 shows the estimated false positive rates, calculated by averaging metric M1 of *PfRecur* and the posterior probability of recrudescence under the CDC model across recurrent isolates. Across the permuted datasets, the CDC model had median false discovery rates of 8.7% (95% confidence interval [CI] 1.2–19.1) in Benguela, 6.5% (95% CI 0.7–18.6) in Lunda Sul and 5.0% (95% CI 1.3–10.2) in Zaire, driven largely by permuted pairs with intermediate posterior probabilities of recrudescence (Supplementary Figure 10). In comparison, *PfRecur* had substantially lower median false discovery rates of 0.6% (95% CI 0.02-7.1) in Benguela, 0.1% (95% CI 0-2.8) in Lunda Sul and 0.4% (0.02-3.9) in Zaire, with false discovery rates remaining comparatively low even as the per-clone marker-wise probability of detection was relaxed from the default value *ω* = 0.9 to *ω* = 0.75 (assuming more clones evade detection tends to augment the posterior probability of recrudescence) (Supplementary Figure 9). The “*≥* 4*/*7” match-counting rule recommended by [16, 17, 25] yielded higher false discovery rates than both *PfRecur* and the CDC model.

**Figure 4.**
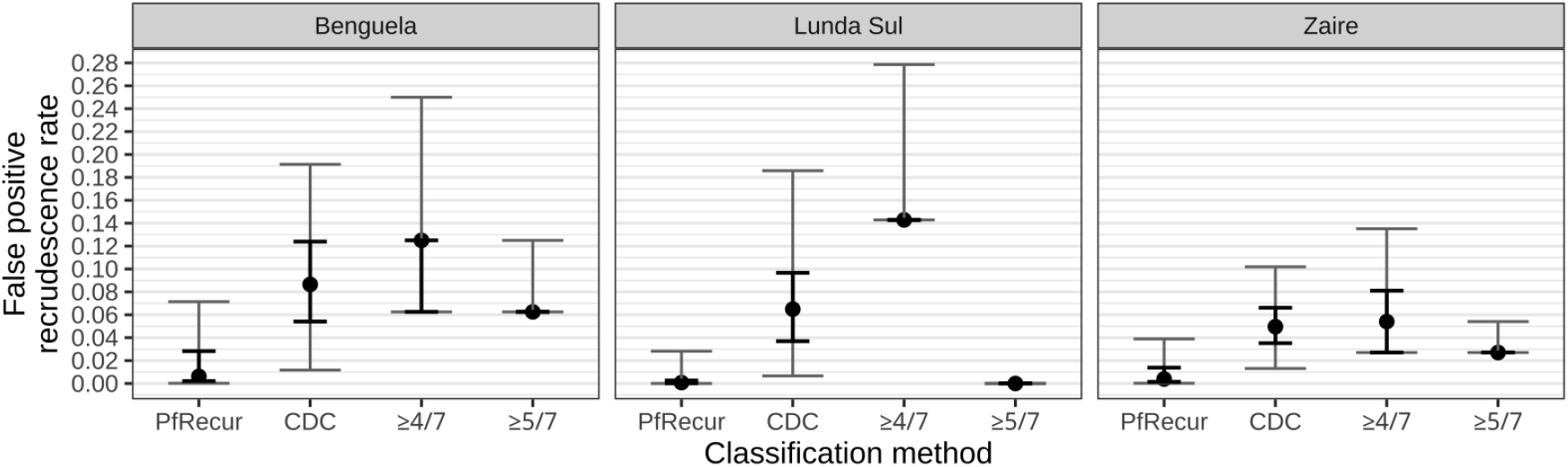
False positive recrudescence rates under *PfRecur* (averaging metric M1) vs the CDC model[8] (averaging the posterior probability of recrudescence) vs a match-counting approach (i.e., treating the presence of one or more shared alleles at 4 or more, or 5 or more, markers as evidence of recrudescence [16, 17, 25]; restricted to permuted pairs with *≥* 1 allele call at each of the 7 markers). Points indicate medians, bold error bars indicate the interquartile range and gray error bars indicate 95% confidence intervals over 500 permuted artificial ‘not-recrudescence’ datasets, each comprising 19 recurrent isolates from Benguela; 13 recurrent isolates from Lunda Sul and 38 recurrent isolates from Zaire.

We note that the false discovery rates are dependent in part on marker diversity: in settings with limited diversity, we would expect *PfRecur* to return the prior distribution over newly-inoculated vs recrudescent clones, and the CDC model to return a 0.5 posterior probability of recrudescence, yielding an elevated apparent false positive recrudescence rate using this method.

## Discussion

Accurate characterisation of ACT failure rates in uncomplicated malaria is essential to guide policies and practices, especially now in Africa where artemisinin resistant *P. falciparum* is spreading and consequently ACTs are under threat [29]. Genotyping of polymorphic alleles has allowed evaluation of antimalarial therapeutic efficacy in malaria endemic areas where study participants are exposed to new infectious bites during follow-up. But in high transmission settings differentiating between reinfection and recrudescence remains a difficult problem. Many malaria infections are polyclonal, posing combinatorial problems since allele counts are not directly observable. The different PCR genotyping methods also have different sensitivities [10]. Limitations of match-counting methods – which do not adjust for multiple comparisons across polyclonal isolates, relative allele frequencies [8, 11] or the imperfect detectability of parasite clones [13, 14] – have been well-established in the literature [14–16]. In addition to having good laboratory techniques for accurate genotype calling, a robust statistical methodology is needed to assess probabilistically whether the recurrent infection is compatible with recrudescence (treatment failure). We show that when infections are polyclonal, match-counting methods and the methodology proposed by the CDC [8] may result in unacceptably high false positive recrudescence rates. In contrast, *PfRecur* has higher specificity whilst retaining good accuracy at identifying recrudescence. When applied to microsatellite data from a recent therapeutic efficacy assessment in Angola [17], our classification model outputs substantially different estimates for one third of the recurrent infections with baseline MOI*>* 1 [8]; however this does not greatly impact efficacy estimates across study arms (Supplementary Table 4). The systematic over-estimation of recrudescence rates is likely to be greater in settings with greater parasite diversity, higher polyclonality, and more frequent reinfection.

The Bayesian model proposed by the CDC has three potential issues. Firstly, a per-clone unobserved allele “penalty” — a multiplicative factor applied to the likelihood when the imputed allele for a clone lies outside the observed set of alleles for a given isolate — is estimated simultaneously during classification (a relatively lax penalty in the range 0.25 to 0.45 was estimated for each study site in [17]). We suggest that this construction increases the estimated probability of recrudescence. This is because the unobserved alleles have little effect on the probability of reinfection (they only affect allele frequency estimates), but tend to augment the chance of allelic matches between the paired isolates and consequently the probability of classifying as a recrudescence. This bias is particularly pronounced for isolates with no genotyping data at a subset of markers, and isolates with unbalanced cardinality across loci. Markers with missing genotypes are included in the estimation procedure, and the per-clone unobserved allele penalty is applied irrespective of whether or not imputed alleles match those in the paired isolate. Isolates with unbalanced cardinality across loci result in a greater chance of introducing unobserved alleles, a feature that may be problematic when MOIs are inflated due to artificially high multiplicities at one marker (e.g. TA109 in the dataset analysed here). Secondly, estimation of the genotyping error appears to be unstable. In the dataset analysed, site specific estimates of genotyping error varied from 0.025 in Lunda Sul to 0.072 in Benguela (i.e. a three fold difference). However, all genotyping was presumably done by the same central laboratory. The difference in estimates suggests possible identifiability issues in the model. As demonstrated by the marker TA109, a major determinant of genotyping error is likely to be the parasite density in the filter paper. Thirdly, the reliability of the output model probabilities depend on convergence of the algorithm. To the best of our knowledge, there has been no in depth study of convergence of this algorithm, and the default parameters suggested (1000 iterations) appear insufficient. In contrast, in our proposed model *PfRecur*, we average analytically over compatible allelic configurations within each isolate to derive an analytically-tractable (discrete) posterior distribution for the number of newly-inoculated vs recrudescent clones within each recurrent isolate that can be evaluated directly, allowing for classification of recurrent isolates with multiplicity of infection up to 9 in the order of seconds.

A limitation of *PfRecur* in low transmission settings is the assumed lack of relatedness structure within or between isolates (the CDC model makes the same assumption). The classifier of Taylor *et al*. [30] for *P. vivax* recurrence explicitly accommodates sibling relationships within isolates (and between isolates, but only in support of relapse), and in addition, includes an optional “fudge-factor” for low-level background relatedness; but the transitive property of relatedness introduces substantial complexity. However, it is shown in [31] that sample allele frequencies, which are plugged into many models of malaria parasite genetic data, can be considered to encode implicitly average relatedness marker-wise.

In conclusion we present an analytically-tractable probabilistic classifier *PfRecur* for estimating recrudescence based on multi-allelic calls in recurrent falciparum malaria. In high transmission settings it is more accurate than current methods of analysis and therefore less likely to overestimate recrudescence rates.

## Methods

### Data from Dimbu *et al*. [17]

We re-analysed data from a six-arm therapeutic efficacy study conducted between February and July in 2021 across 3 provinces in Angola [17]. Outpatients presenting to urban clinics situated in provincial capitals were screened for inclusion in the study. Enrolment criteria included uncomplicated *P. falciparum* monoinfection and either a history of fever or an axillary temperature reading *≥* 37.5^*°*^C. To avoid confounding due to transmission intensities, some inclusion criteria varied by province. In Benguela (low to moderate transmission), children between 6 and 143 months of age with 1,000—100,000 parasites/*µ*L at baseline were enrolled. In Lunda Sul (moderate to high transmission) and Zaire (moderate to very high transmission), children were enrolled with a narrower age range of 6 to 59 months, and higher baseline parasitemias of 2,000—200,000 parasites/*µ*L. Parasitemia was quantified using microscopy. 622 patients were enrolled across the study arms.

Study participants were treated with 3-day regimens of either artemether-lumefantrine or artesunate-amodiaquine (Lunda Sul and Zaire), and dihydroartemsinin-piperaquine or artesunate-pyronaridine (Benguela). Dosing was determined by weight bands in accordance with manufacturer guidelines. Antimalarial treatment was largely supervised. Follow-up, entailing clinical examination and slide microscopy, occurred on days 1, 2, 3, 7, 14, 21 and 28 in addition to days 35 and 42 for patients treated with dihydroartemsinin-piperaquine and artesunate-pyronaridine. Recurrent infections, characterised by microscopy-detected asexual *P. falciparum* parasitemia from day 7 to the end of follow-up, were identified in 71 patients.

Genotyping was performed for 70 pairs of baseline and recurrent isolates, and an additional 42 baseline isolates. DNA was extracted from dried blood spots and *P. falciparum* diagnosis was confirmed by PCR. For a panel of 7 neutral microsatellite markers (*M2490, M313, M383, PfPK2, POLYA, TA1, TA109*), fragment lengths were then assessed using capillary electrophoresis. Classification of reinfection vs recrudescence was performed using the Bayesian CDC model [8], our novel probabilistic approach, *PfRecur*, and simple match-counting algorithms (at least 4/7 [16, 17, 25] or 5/7 matches, where a per-marker observation is a match if some alleles are the same at enrolment and recurrence), stratified by study site.

### Marker cardinality as a function of parasitemia in [17]

We explored the relationship between MOI (defined as the maximum cardinality across the genotyped panel of 7 neutral microsatellite panel) and parasitemia for 181 isolates with genotyping data available, using the Jonckheere-Terpstra test [32], with *p*-values derived under a normal approximation using the R function PMCMRplus::jonckheereTest [33]. We also examined the within-host diversity of each marker, as a function of parasitemia. We defined the within-host diversity of each marker for each isolate as the complement of Nei’s gene identity metric [34], taking a uniform distribution over compatible allelic configurations at that locus (i.e., all possible ways of allocating alleles observed at that locus to the number of clones given by the MOI). Nei’s gene identity metric is formulated as the sum of squares of allele frequencies; the complement can be interpreted as the probability of differing alleles when a pair of clones is sampled (with replacement) from the isolate (see Appendix C.1).

### PfRecur

*PfRecur* classifies a recurrent infection as either a recrudescence or a reinfection based on either the posterior probability of there being at least one recrudescent clone (M1); or the posterior expected proportion of recrudescent clones (M2) (Figure 5). A complete description of the framework is provided in Appendix A; below, we provide a brief outline, using the terms marker and locus interchangeably. Classification of recurrent infections is done for each pair in a group of patients, where the grouping is determined by the user (usually by study or study site, depending on the context). The model is not fully Bayesian in that there is no way of specifying multilevel groupings.

**Figure 5.**
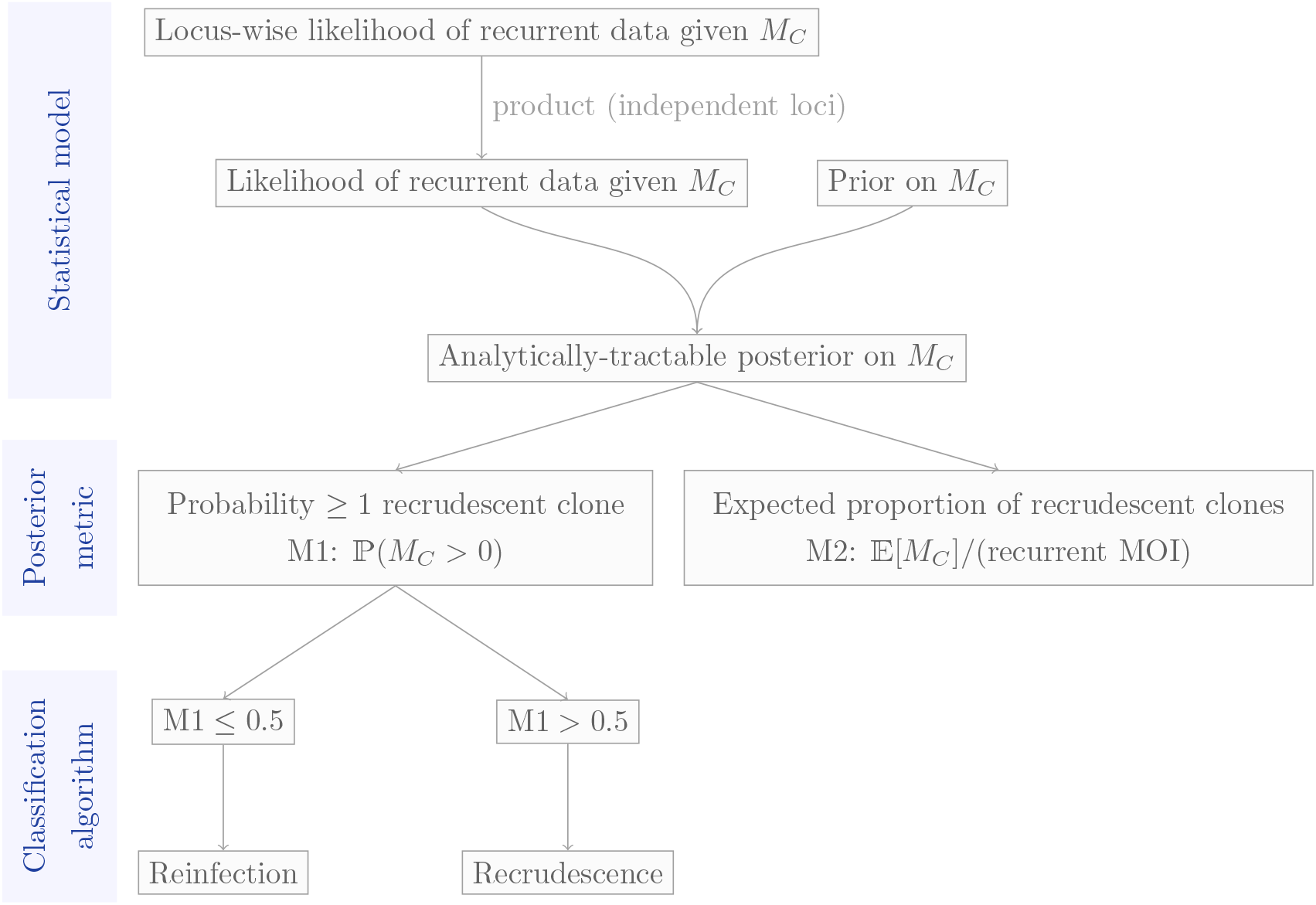
*PfRecur* is designed to classify a recurrence as either a recrudescence or a reinfection using a posterior summary of *M*_*C*_, the number of recrudescent clones in the *r*th isolate.

#### Box 1

*PfRecur* model assumptions

- **Unlinked loci**: valid if loci lie on different chromosomes (because chromosomes assort independently in meiosis) or inter-locus distances are large (because then loci are more likely separated by one or more recombination break points in meiosis).
- **Clones within isolates are independent as are non-recrudescent clones between isolates**: violated by the presence of sibling parasites within isolates [30]; additionally violated if the average population-level relatedness is high (although baseline isolates 1, …, *n* partially encode average relatedness [31]) and/or if the population is structured (e.g., by geographic barriers in study site, or by household effects among study participants).
- **Uniform distribution of allelic states for each isolate** (i.e., any configuration of alleles compatible with the observed MOI and the set of genotypes observed at a given locus is modelled to be equally likely): enforced in the absence of quantitative data on relative allelic abundance.
- **Reinfection and recrudescence are not mutually-exclusive**: the recurrent isolate *r* comprises a mixture of recrudescent clones drawn from the paired baseline isolate *b*, and newly-inoculated clones drawn from the contemporaneous population at large, which is approximated by the remaining baseline isolates 1, …, *n*.
- **Non-parametric genotyping error**: we accommodate non-parametric genotyping error in baseline isolates *relative* to the recurrent isolate, specified by the probability that each allele called in a baseline isolate matches an allele called in the recurrent isolate *r*.
- **No undetected clones in the baseline isolates** 1, …, *n*: we assume that the consequences of ungenotyped clones (if any) ‘average out’ over isolates 1, …, *n* from which baseline population allele frequencies are derived.
- **Ungenotyped clones in the paired baseline isolate** *b*: the allelic states of undetected clones in the paired baseline isolate are imputed using allele frequencies derived over the unpaired baseline isolates 1, …, *n*, with a truncated binomial model for the number of clones genotyped per locus.
- **Ungenotyped clones in the recurrent isolate** *r*: observed alleles in the recurrent isolate are allocated over successfully genotyped clones only, with a truncated multinomial model for the number of clones genotyped per locus.

### Overview of the statistical model

*PfRecur* classifies infections based on an analytically tractable Bayesian model constructed around the set of assumptions detailed in Box 1. Under the model, each isolate is treated as a set of genetically-distinct clones; the cardinality of this set is referred to hereafter as the multiplicity of infection (MOI). For each isolate, we observe a set of alleles *G*_*𝓁*_ at loci *𝓁* ∈ *{*1, …, *L}*, with locus-wise cardinality *M*_*𝓁*_ = |*G*_*𝓁*_|. We take the MOI to be the maximum cardinality over all loci: *M* = max_1≤*𝓁*≤*L*_ *M*_*𝓁*_. Given bulk genotypic data with no quantitative information on the intra-isolate abundance of each allele^1^, we assume a uniform distribution over the sets of allelic configurations that are compatible with *G*_*𝓁*_. We assume independence between clones within isolates, between non-recrudescent clones across isolates, and between loci.

The indices *r, b* and *i* are used to distinguish isolates that are handled differently under the model. Index *r* corresponds to the recurrent parasite isolate for which classification is being performed; index *b* corresponds to the paired baseline isolate (i.e., it is from the same study participant as isolate *r*). Index *i* = 1, …, *n* iterates over *n* baseline isolates that are not paired with isolate *r*: they are from different study participants.

The recurrent isolate *r* (of MOI *M* ^(*r*)^) is modelled as a mixture of *M*_*C*_ recrudescent clones and *M*_*I*_ = *M* ^(*r*)^ − *M*_*C*_ newly-inoculated clones. For each recurrent isolate *r*, the target of inference under the *PfRecur* statistical model is the posterior distribution over *M*_*C*_, with state space *{*0, …, *M* ^(*r*)^*}*:

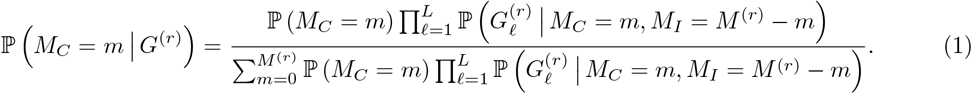

Figure 6 depicts the model structure. The likelihood of the locus-wise recurrent data, 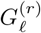, is conditional on the random variables *M*_*C*_ and *M*_*I*_ = *M* ^(*r*)^ − *M*_*C*_. It is also conditional on the locus-wise baseline data, 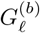 and 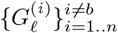, and on the recurrent and baseline MOIs, which are derived from the complete recurrent and baseline data. Although these variables are data derived, they are not treated as random variables under the model. The likelihood also features other user-defined inputs that are not treated as random variables. These include a genotyping detection rate, *ω*, and a locus-wise genotyping error matrix, *δ*_*𝓁*_. The prior on *M*_*C*_ is conditional on *M* ^(*r*)^ and on a prior parameter *β*. There is no prior on *M*_*I*_ in Equation (1) since *M*_*I*_ is deterministic given *M* ^(*r*)^ and a realisation of *M*_*C*_. Thanks to an analytically tractable likelihood (next section), Equation (1) is analytically tractable. As such, inference under the *PfRecur* framework does not require a numerical sampler or an optimisation algorithm.

**Figure 6.**
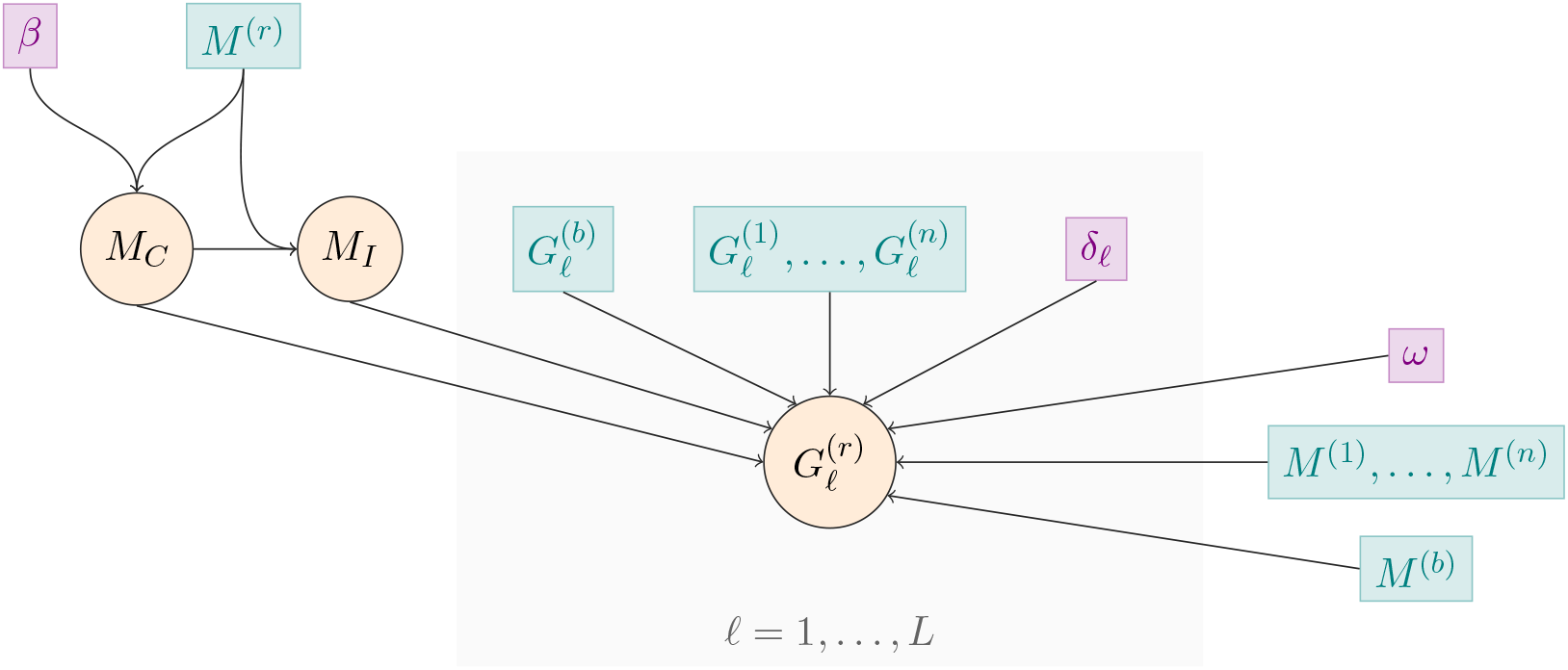
A graphical representation of the Bayesian statistical model within the *PfRecur* framework. Random variables are circled. Data driven quantities are shown in teal; user-specified values are shown in violet.

### Model likelihood

We derive multiple expressions in Appendix A (see Equations (24), (26), (27) and (28)), which together can be used to evaluate the likelihood:

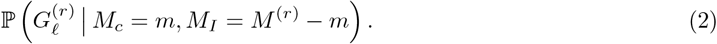

The various steps used to conceptualize the likelihood are sketched out below. Throughout, newly-inoculated clones are drawn from population *I* (the contemporaneous population at large), which is approximated under the model by the baseline isolates *i* = 1, …, *n*. The recrudescent clones are drawn from population *C*, comprised of clones in the paired baseline isolate *b*. In an intermediate step, the probability of 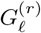 is modelled conditional on allele frequencies, whereby the probability that a clone drawn from population *S* ∈ *{I, C}* harbours allele *α* at locus *𝓁* is equated to the population allele frequency 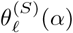. In later steps, allele counts supersede allele frequencies.

### Modelling the number of clones genotyped per locus

In the allelic data of a given isolate, some loci have cardinalities lower than the MOI. The lower cardinality could occur because multiple clones within the isolate share identical alleles at this locus, because the MOI is supuriously elevated by genotying errors, or because some clones are not genotyped at this locus. We model the number of genotyped clones per locus in the recurrent isolate *r* at loci with cardinalities strictly less than the observed MOI (i.e. 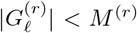), thereby allowing some clones in the recurrent isolate to evade detection at some loci (multiple clones sharing identical isolates is also addressed — see next section). A spuriously elevated MOI is not accounted for.

We construct a model of the number of clones genotyped per locus in the recurrent isolate *r*, i.e., the number of clones that contribute to the observation 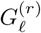, as follows. Each clone in isolate *r*, irrespective of whether it is drawn from population *I* or *C*, is detected with probability *ω* at locus *𝓁*. Then, given *M*_*C*_ = *m* and *M*_*I*_ = *M* ^(*r*)^ − *m*, the number of detected clones 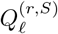 derived from populations *S* ∈ *{I, C}* at locus *𝓁* follow the truncated multinomial distribution

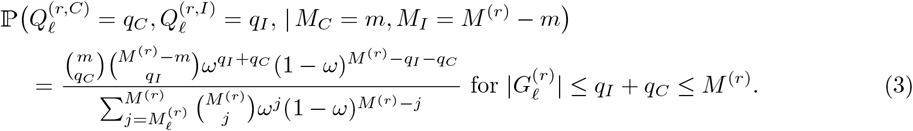

### Allocating observed alleles to genotyped clones

We average analytically over compatible allelic configurations in isolate *r* using the inclusion-exclusion principle. Suppose there are 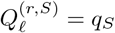 genotyped clones drawn from populations *S* ∈ *{I, C}* at locus *𝓁* in isolate *r*. Then the probability of observing the set of alleles 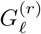 at locus *𝓁* takes the form

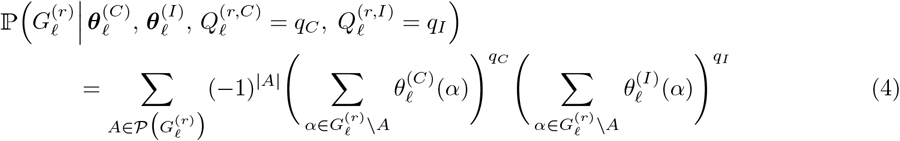

where 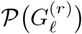 denotes the power set of 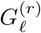; that is, the set of all subsets of 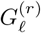 encompassing the empty set. Convolving Equation (4) over the locus-wise truncated multinomial model of genotyped clones (3) yields the probability of the observation 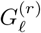 at locus *𝓁* for the recurrent isolate *r*, formulated with respect to the population allele frequencies 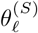, *S* ∈ *{C, I}*.

### Modelling allele frequencies

The derivation of population allele frequencies under our model is two-fold. Denote by *H*_*𝓁*_ the set of possible alleles at locus *𝓁* (equipped with an arbitrary ordering) and by 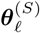 the vector of population allele frequencies over *H*_*𝓁*_. We begin by deriving allele frequencies 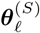 conditional on the vector of per-isolate allele counts **C**_*𝓁*_ for each baseline isolate *b* or 1, …, *n* over *H*_*𝓁*_; and later address the distribution of per-isolate allele counts **C**_*𝓁*_, which are not directly observed using bulk genotypic data.

For population *I*, we derive allele frequencies under a Bayesian multinomial-Dirichlet model of baseline isolates *i* = 1, …, *n* (which excludes the paired baseline isolate *b*). Allele frequencies are formulated over *clones* in the baseline isolates *i* = 1, …, *n*, whereby each isolate is effectively weighted by its MOI (i.e., high MOI isolates are more informative). Under the assumption of clone-wise independence (both within and across isolates), we obtain the multinomial likelihood

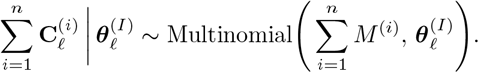

Taking a uniform prior for 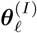 over the |*H*_*𝓁*_| − 1 simplex yields the posterior

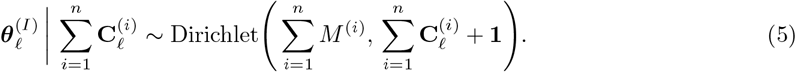

For population *C*, allele frequencies are largely informed by the paired baseline isolate *b*, but not exclusively because the alleles of ungenotyped clones in isolate *b* are imputed using population *I* allele frequencies, which are based on isolates *i* = 1, …, *n* (Equation (5)). This imputation explains the omission of isolate *b* in the formulation of allele frequencies for population *I*. Akin to the model of genotyped clones for isolate *r* (Equation (3)), the number of genotyped clones 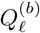 in isolate *b* that contribute to the observation 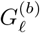 is modelled to follow a truncated binomial distribution, with support 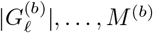 and success probability *ω*. Given 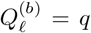, we denote by 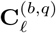 the vector of allele counts over the *q* clones in isolate *b* that are genotyped at locus *𝓁*. We model 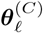 as a deterministic function of 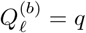, the per-isolate allele counts 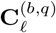 of the genotyped clones in isolate *b*, and 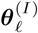,

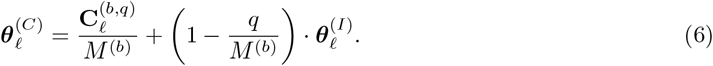

### Modelling genotyping errors

We account for genotyping error when modelling the per-isolate baseline allele counts 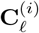 and 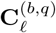. Genotyping errors are modelled using a user-specified right-stochasic error matrix *δ*_*𝓁*_ for each locus *𝓁*, with the interpretation that *δ*_*𝓁*_(*α, α*^*′*^) yields the probability that an allele called as *α* in a baseline isolate *b* or *i* = 1, …, *n* matches an allele called as *α*^*′*^ in a recurrent isolate *r* at locus *𝓁*. The *PfRecur* framework is marker-agnostic, in that the non-parametric error model *δ*_*𝓁*_ can be adapted to different marker types. In the present study, we consider a normalised geometric model adapted to length polymorphic microsatellite markers (akin to [8]), parametrised by a genotyping error probability *ε* where, for allele lengths *i, j*

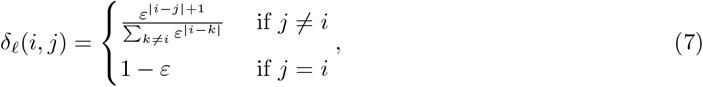

where the sum in the denominator is taken over the allelic lengths in the set *H*_*𝓁*_.

### Deriving moments of allele counts

The locus-wise probability (4) of observed genotypes for recurrent isolate *r* is formulated as a multinomial expression of the population allele frequencies 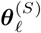. Convolving Equation (4) over the modelled distributions of 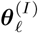 (5) and 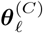 (6), which are conditioned on per-isolate allele counts, yields a multinomial expression of baseline allele counts (Equation (19)).

Because individual clones and the alleles they carry are not directly observable in multiclonal isolates genotyped using standard methods (e.g., not single-cell genotyping), allele counts for multiclonal isolates must be derived under an appropriate model [20–24]. By Jensen’s inequality, the expected per-isolate allele counts 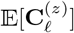 (which are straightforward to compute) cannot be directly substituted for 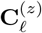 in Equation (19). Convolving the locus-wise probability (19) over compatible allelic configurations in baseline isolates *b* and *i* = 1, …, *n* requires calculating moments of the per-isolate allele counts

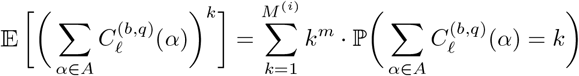

and the sample allele counts (i.e., per-isolate allele counts summed over one or more baseline isolates)

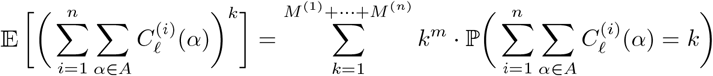

aggregated over allelic subsets *A* ⊆ *H*_*𝓁*_ (Appendix A.2).

In Appendix A.3, we derive these moments from first principles under our framework. In brief, we use a simple combinatorial argument based on ordered partitions to derive probability mass functions and consequently cumulants of the per-isolate allele counts. To adjust per-isolate allele counts for genotyping error, we adopt a Poisson binomial model: the number of distinct alleles in a given set *A* ⊂ *H*_*𝓁*_ that are harboured by clones in a baseline isolate is modelled as a sum of |*G*_*𝓁*_| independent, but not identically distributed, Bernoulli random variables with respective success probabilities 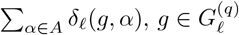.

To compute moments of the sample allele counts (under the assumed independence of clones within and between baseline isolates), we first sum cumulants of the per-isolate allele counts to recover cumulants of the sample allele counts. We then exploit complete exponential Bell polynomials to map cumulants of the sample allele counts to moments of the sample allele counts, as required. We adopt this construction because the order of these moments (up to *M* ^(*r*)^) is likely, in practical settings, to be smaller than the number of baseline isolates *n* over which allele counts are aggregated.

### Prior of the statistical model

Since mixtures of newly-inoculated and recrudescent clones are comparatively unlikely in low transmission transmission settings, we take a symmetric prior, which weights pure reinfections and pure recrudescences more heavily than intermediate mixtures. We implement this through a symmetric beta binomial distribution

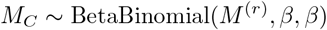

with 0 *< β* ≤ 1. In the case *β* = 1, we recover a uniform distribution over the breakdown of newly-inoculated vs recrudescent clones. In the limit *β* → 0, the prior probability of all intermediate mixtures approaches zero, whereby reinfection and recrudescence constitute mutually exclusive categories.

### Posterior metrics

We generate two posterior metrics for each recurrence: the posterior probability of at least one recrude-scent clone

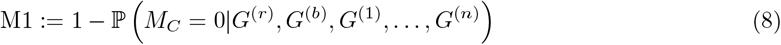

and the posterior expected proportion of recrudescent clones

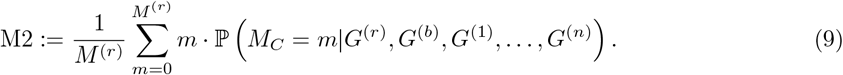

### Application of posterior metrics

For conceptual consistency with the CDC model [8], we perform classification using metric M1 of *PfRecur* : a recurrent isolate *r* is classified as a recrudescence if M1 *>* 0.5, and as a reinfection otherwise. Down-stream efficacy estimates are computed using metric M1, rather than dichotomised classifications, in line with recommendations from the CDC [8, 17]. Metric M2 serves as a supplementary descriptor for recurrences comprising a mixture of newly-inoculated and recrudescent clones, and is used to validate the model against simulated data for which these mixtures are known (see below).

### R software

*PfRecur* is implemented as an R package (available at https://github.com/somyamehra/PfRecur). This largely relies on base R functionality [35], with additional dependencies on `copula::Stirling2` and `copula::Stirling1` [36] (to evaluate Stirling numbers of the second and unsigned first kind respectively); `PDQutils::cumulant2moment` and `PDQutils::moment2cumulant` [37] (to map between cumulants and moments respectively); `poisbinom::dpoisbinom` [38] (to evaluate the density function of the Poisson binomial distribution); and `VGAM::dbetabinom.ab` [39] (to evaluate the density function of the beta binomial prior). The package accommodates isolates of MOI 9 and fewer (to avoid numerical instability when evaluating the posterior).

As input, the *PfRecur* package requires categorical (presence/absence) genotypic data in a list of binary matrices, where each matrix corresponds to a marker; named matrix columns correspond to alleles; named matrix rows correspond to isolates; and matrix elements are set to 1 if the corresponding allele has been detected in the relevant isolate, and 0 otherwise. Additional user-specified parameters include the per-clone marker-wise probability of detection *ω*, and a list of marker-wise row-stochastic genotyping error matrices *δ*_*𝓁*_.

Given a recurrent isolate *r*, paired baseline isolate *b*, and baseline isolates 1, …, *n*, the function `PfRecur::-evaluate_posterior` returns the discrete posterior distribution for *M*_*C*_ over the state space *{*0, …, *M* ^(*r*)^*}* in addition to metrics M1 (8) and M2 (9), in the form of a named list.

### Simulation study

To validate *PfRecur*, we simulate recurrent isolates as mixtures of newly-inoculated and recrudescent clones (Appendix B). In brief, we consider isolates with MOIs up to 9 (mean baseline MOI ≈ 3), genotyped at 7 unlinked multi-allelic markers (each with between 10 and 30 distinct alleles). We permit siblings within isolates, violating the assumed independence of clones under *PfRecur*. Each simulated clone is detected at each marker with probability *ω* = 0.9. Genotyping error is applied to the set of alleles harboured by detected clones in each baseline isolate with probability *ε* = 0.05, in accordance with the length-dependent normalised geometric model (7). We simulate 40 baseline datasets, each comprising 25 isolates. For a baseline isolate of MOI *M* ^(*b*)^, we simulate paired recurrences with MOI *M* ^(*r*)^ = 1, …, 9 and *m* = 0, …, min*{M* ^(*b*)^, *M* ^(*r*)^*}* recrudescent clones. We apply our probabilistic classifier *PfRecur* to recover the posterior distribution for the number of newly-inoculated vs recrudescent clones within each simulated recurrent isolate under a uniform prior (*β* = 1); the underlying parameters *ω* and *ε* are assumed to be known. The results in the main text, pertaining to metrics M1 and M2, are aggregated across 28773 simulated recurrences.

### Reanalysis of Dimbu *et al*. [17]

Using *PfRecur* we perform a re-analysis of [17], with baseline isolates stratified by study site (whereby allele frequencies for newly-inoculated clones are modelled to be site-specific). By default, we set *β* = 0.25 for the prior; *ω* = 0.9 for the per-clone marker-wise probability of detection in each baseline/recurrent pair; and *ε* = 0.05 for the genotyping error probability, under the normalised geometric model (7). We additionally perform a sensitivity analysis for *ω* ∈ [0.75, 1] and *ε* ∈ [0, 0.25]. Classification is performed with both the entire 7 microsatellite marker set, and also omitting the *TA109* marker.

We compare posterior metric M1 of *PfRecur* against the “gold-standard” CDC model [8], which was originally used to analyse [17]. In the present study, we have re-run code provided openly by Plucinksi and colleagues [40] (with 100,000 iterations for the Gibbs sampler); posterior probabilities based on all 7 microsatellite markers may differ from those reported in [17] due to the stochastic nature of the MCMC algorithm.

### False positive recrudescence rates

To estimate false positive rates for calling recrudescence, we generate 500 artificial ‘non-recrudescence’ datasets from [17] by generating random derangements of baseline study participants labels within each study site, whereby permuted baseline/recurrent pairs cannot be derived from the same individual and therefore cannot represent recrudescences. The generation of permuted datasets, rather than permuted pairs, is necessitated by the construction of the CDC model [8]. We perform classification for these permuted datasets using both *PfRecur* (metric M1) and the CDC model [8, 40] (with 10,000 iterations for the Gibbs sampler due to computational time constraints). For each permuted dataset, we compute the false positive recrudescence rate by averaging the posterior probability of recrudescence (under the CDC model) or metric M1 (under *PfRecur*) over recurrent isolates. In addition, we consider a match-counting approach, treating the presence of one or more shared alleles at least 4 or 5 markers as evidence of recrudescence. For the panel of 7 neutral microsatellites, no WHO-endorsed guidelines are available but [16, 17, 25] support the “*≥* 4*/*7 rule”^2^. To avoid ambiguity, we restrict match-counting classification to baseline/recurrent pairs with at least one allele call at each of the 7 markers.

## Data Availability

The *PfRecur* framework has been implemented in an eponymous R package, openly available in a GitHub repository: https://github.com/somyamehra/PfRecur.
This study uses open access data that has previously been published by Dimbu et al (2024) (DOI:10.1128/aac.01525-23). Parasite densities and clinical metadata have been retrieved from Supplemental Table S4 of Dimbu et al (2024), while genotypic data have been retrieved from an accompanying GitHub repository (https://github.com/MateuszPlucinski/AngolaTES2021).
We have re-run code provided openly by Dr Mateusz Plucinski and colleagues (with very minor input/output modifications), which implements the model detailed in Plucinski et al (2015) (DOI:10.1128/AAC.00072-15) and is available in a GitHub repository: https://github.com/MateuszPlucinski/AngolaTES2021.
For completeness, all code and data relevant to this study (including the data of Dimbu et al (2024) and the implementation of Plucinski et al (2015)) have been collated in a GitHub repository: https://github.com/somyamehra/PfTreatmentFailure

https://github.com/somyamehra/PfTreatmentFailure

https://github.com/somyamehra/PfRecur

## Code and data availability

The *PfRecur* framework has been implemented in an eponymous R package, openly available in a GitHub repository: https://github.com/somyamehra/PfRecur.

This study uses open access data that has previously been published by Dimbu *et al*. [17]. Parasite densities and clinical metadata have been retrieved from Supplemental Table S4 of Dimbu *et al*. [17], while genotypic data have been retrieved from an accompanying GitHub repository [40].

We have re-run code provided openly by Dr Mateusz Plucinski and colleagues (with very minor input/output modifications), which implements the model detailed in Plucinski *et al*. [8] and is available in a GitHub repository [40]: https://github.com/MateuszPlucinski/AngolaTES2021.

For completeness, all code and data relevant to this study (including the data of Dimbu *et al*. [17] and the implementation of Plucinski *et al*. [8]) have been collated in a GitHub repository: https://github.com/somyamehra/PfTreatmentFailure.

## Acknowledgements

We thank Dr Mateusz Plucinski from Centers for Disease Control and Prevention, Atlanta for thoughtful and constructive criticism of the manuscript.

We thank the authors of Dimbu *et al*. [17] for making their code and data openly available. JAW is a Sir Henry Dale Fellow funded by the Wellcome Trust (223253/Z/21/Z). NJW is a Principal Research Fellow funded by the Wellcome Trust (093956/Z/10/C). A CC BY or equivalent licence is applied to the author accepted manuscript arising from this submission, in accordance with the grant’s open access conditions. ART is a Marie Sklodowska-Curie Fellow (project number 101110393) funded by the European Union. Views and opinions expressed are however those of the author(s) only and do not necessarily reflect those of the European Union or granting authority. Neither the European Union nor the granting authority can be held responsible for them.

## Author contributions

Methodology: S.M. and J.A.W. Formal analysis, visualisation: S.M. Writing (original draft): S.M., A.R.T., N.J.W and J.A.W. Writing (review and editing): S.M., A.R.T., M.I., N.J.W. and J.A.W. Conceptualisation: N.J.W. and M.I. Supervision: N.J.W. and J.A.W.

## Competing interests

The authors have no competing interests to declare.

## A Derivation of model likelihood

We model an isolate *r* from a recurrent infection as a mixture of clones drawn from two populations: population *C* comprising clones in the paired baseline isolate *b* (this represents reCrudescence); and population *I*, reflecting the contemporaneous at large but approximated by the baseline population for which baseline isolates 1, …, *n* (excluding the baseline isolate *b*, which is paired with *r*) constitute a finite sample (this represents reInfection).

Under our model, the probability that a given clone in isolate *r* exhibits allele *α* is equated to the frequency of *α* in the population from which it is derived. For population *I*, allele frequencies are derived across clones in isolates 1, …, *n* under a multinomial-Dirichlet model (with a uniform prior over the standard simplex); for population *C*, we instead consider allele frequencies across clones in isolate *b*. We make the simplifying assumptions of marker-wise independence (i.e., we assume that loci are unlinked); neutral markers; and clone-wise independence, both within isolates and between non-recrudescent clones across isolates (i.e., we ignore relatedness, which amplifies allelic sharing between closely-related clones [30]).

The principal difficulty arises because per-isolate genotypic data are generated in bulk. We observe the presence or absence of detection of an allele within an isolate, but not the allocation of alleles to individual clones, and thus not the per-isolate allele counts. Here, we take a uniform distribution over all allelic configurations that are compatible with the set of observed alleles. Further complications include imperfect detectability of clones and inaccurate genotyping. Accordingly, we account for undetected clones in the paired baseline *b* and recurrent *r* isolates, under a marker-wise truncated binomial model. The locus-wise probability of observations for isolate *r* is formulated over successfully genotyped clones only. In contrast, allelic states for undetected clones in isolate *b* are imputed based on population *I* allele frequencies (this imputation is the rationale for excluding isolate *b* from population *I*). We additionally allow for genotyping error in baseline isolates *b* and 1, …, *n* relative to the recurrent isolate *r*, specified by the probability that an allele called in a baseline isolate matches an allele called in isolate *r*.

In the following sections, we derive an analytic locus-wise likelihood of observed genotypes for isolate *r* that addresses these complications systematically. We first formulate the locus-wise probability of observations for the recurrent isolate *r* with respect to allele frequencies in populations *I* and *C* (Section A.1), followed by per-isolate allele counts across clones in baseline isolates *b* and 1, …, *n* (Section A.2). Characterising per-isolate allele counts from categorical (presence/absence) data on multiclonal isolates is a non-trivial problem that has been addressed previously [20–24], which are all reviewed in [41]. However, to evaluate the likelihood, we note that it is sufficient to compute *moments* of allele counts summed over one or more baseline isolates. This is a fundamentally easier problem, and permits an explicit solution under our simplifying assumptions (Section A.3).

## Notation

We begin by introducing notation and several additional assumptions. We consider isolates that have been genotyped at a panel of *L* markers. At locus *𝓁* ∈ *{*1, …, *L}*, for each isolate *z*, we observe a set of alleles 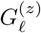. We denote by 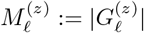 the observed cardinality (i.e., the number of distinct alleles) at locus *𝓁* for isolate *z*. The MOI for isolate *z* is taken to be the maximum observed cardinality across the set of *L* loci, that is,

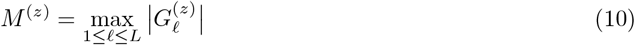

(this assumption can be readily relaxed).

We thus assume that isolate *z* comprises a composite of *M* ^(*z*)^ clones. We index these clones *j* (arbitrary ordering), and denote by 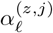 the allele carried by the *j*^th^ clone in the *z*^th^ isolate at locus *𝓁*. If *M* ^(*z*)^ *>* 1 we do not observe 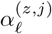. Assuming perfect detectability across clones and perfect genotyping accuracy, we can at most observe whether or not at least one clone in isolate *z* carries allele *α* at locus *𝓁*, that is,

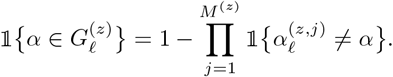

However, we do not observe *how many* or which (under the arbitrary ordering) of the *M* ^(*z*)^ clones within isolate *z* carry allele *α* at locus *𝓁*.

We model genotyping error for baseline isolates as follows. Denote by 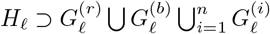 the set of known alleles at locus *𝓁*. We say that an allele identified as *α* ∈ *H*_*𝓁*_ in a baseline isolate *b* or 1, …, *n* matches allele *α*^*′*^ ∈ *H*_*𝓁*_ in isolate *r* with probability *δ*_*𝓁*_(*α, α*^*′*^) *≥* 0 where

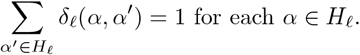

For notational convenience, we impose an arbitrary ordering on elements in the set *H*_*𝓁*_; the notation ***θ***_*𝓁*_ is used to denote a vector of allele frequencies, while **C**_*𝓁*_ is used to denote a vector of per-isolate allele counts across the ordered set *H*_*𝓁*_. *δ*_*𝓁*_ can then be formulated as a right stochastic matrix (i.e.,with rows summing to 1). A summary of notation is provided in Table 2.

**Table 2.** Summary of notation.

|  | Quantity | Interpretation |
| --- | --- | --- |
| *Observed* | $H_\ell$ | Set of possible alleles at locus $\ell$ |
| | $G_\ell^{(z)}$ | Set of alleles observed for isolate $z$ at locus $\ell$ |
| | $M_\ell^{(z)} = \vert G_\ell^{(z)}\vert $ | Cardinality at locus $\ell$ for isolate $z$ |
| | $M^{(z)}$ | Multiplicity of infection for isolate $z$ |
| *User-specified* | $\omega$ | Per-clone locus-wise probability of being detected at locus $\ell$ (applied to clones in isolates $b$ and $r$ only) |
| | $\delta_\ell(\alpha, \alpha')$ | Model of genotyping error: probability that an allele identified as $\alpha$ in a baseline isolate $b$ or $i = 1, \dots, n$ matches allele $\alpha'$ in a recurrent isolate $r$ locus $\ell$ |
| *Estimated* | $M_S$ | Number of clones in isolate $r$ derived from population $S \in \{I, C\}$ |
| *Unobserved* | $\alpha_\ell^{(z,j)}$ | True allele harboured by clone $j$ of isolate $z$ at locus $\ell$ |
| *Modelled* | $\theta_\ell^{(P)}(\alpha)$ | Frequency of allele $\alpha$ at locus $\ell$ in population $P \in \{I, C\}$ |
| | $Q_\ell^{(r,S)}$ | Number of clones in isolate $r$ that are derived from population $S \in \{I, C\}$ and have been detected at locus $\ell$ |
| | $Q_\ell^{(b)}$ | Number of clones in isolate $b$ that have been detected at locus $\ell$ |
| | $C_\ell^{(i)}(\alpha)$ | Count of allele $\alpha$ across clones in isolate $i = 1, \dots, n$ at locus $\ell$ |
| | $C_\ell^{(b,q)}(\alpha)$ | Count of allele $\alpha$ across the $q$ clones in isolate $b$ that have been genotyped (in)correctly at locus $\ell$ |
| | $X_\ell^{(z)}(A)$ | Adjusting for genotyping error relative to isolate $r$, the number of alleles observed for baseline isolate $z = b$ or $z = 1, \dots, n$ at locus $\ell$ that lie in the set $A \subset H_\ell$ |
| *Other* | $s(m, k)$ | Unsigned Stirling number of the first kind |
| | $S(m, k)$ | Stirling number of the second kind |
| | $\mathcal{P}(X)$ | Power set of $X$, i.e., the set of all subsets of $X$ encompassing the empty set |

### A.1 A locus-wise model predicated on allele frequencies

We begin by formulating a locus-wise model of observed alleles 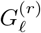 for isolate *r*, with respect to allele frequencies in populations *I* and *C*. The derivation of allele frequencies is deferred to Section A.2.

To accommodate incomplete detection of clones in the recurrent isolate *r*, we allocate observed alleles to successfully genotyped clones only. We adopt a locus-wise truncated multinomial model for the number of successfully genotyped clones at each locus *𝓁*. We assume that each clone in isolate *r*, whether recrudescent or newly-inoculated, is successfully genotyped at locus *𝓁* with equal probability *ω*. Suppose the recurrent isolate *r* comprises *M*_*C*_ = *m* clones from population *C* and *M*_*I*_ = *M* ^(*r*)^ − *m* clones from population *I*. Then the numbers of clones 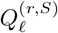 in isolate *r* that are derived from populations *S* ∈ *{C, I}* and successfully genotyped at locus *𝓁* (that is, contribute to the observation 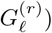) take the form

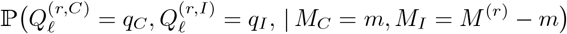

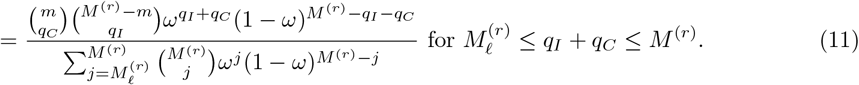

Suppose 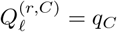 clones from population *C* and 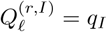 clones from population *I* are successfully genotyped at locus *𝓁* in isolate *r*. As in [23, 42], we use the inclusion-exclusion principle to average over the different ways to allocate alleles in 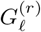 to *q*_*C*_ +*q*_*I*_ clones; the inclusion-exclusion principle circumvents the need to allocate alleles to clones explicitly. Let 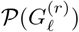 denote the power set of 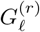, that is, the set of all subsets of 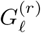 (encompassing the empty set). Then the conditional probability of the observation 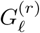 at locus *𝓁* is

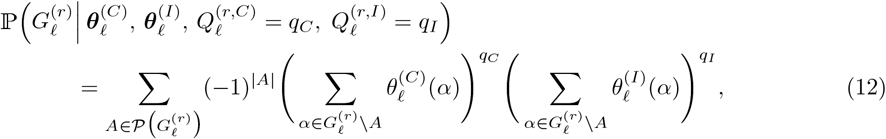

where 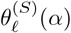 is the frequency of allele *α* at locus *𝓁* in population *S* ∈ *{I, C}*.

Convolving Equation (12) the distribution of successfully genotyped clones (11) then yields

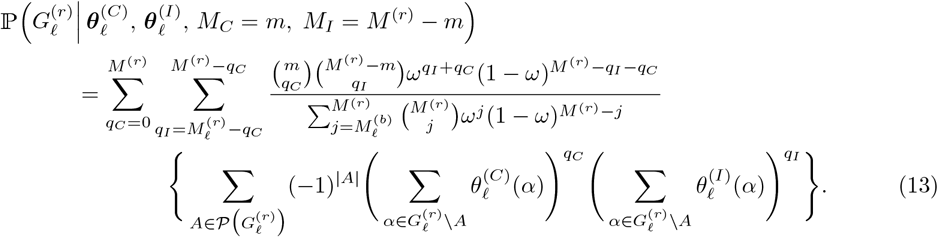

***Remark*** : Rather than modelling (un)genotyped clones in recurrent isolate *r* directly, we could have taken a uniform distribution over all allelic configurations of *M* ^(*r*)^ clones such that each allele in the observed set 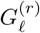 is carried by at least one clone at locus *𝓁*; this would implicitly allow for the incomplete detection of clones in isolate *r*. Under this construction, the conditional probability of the observation 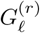 at locus *𝓁* would take the form

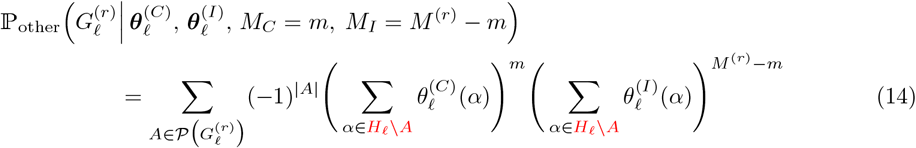

with differences to Equation (12) highlighted in red. We have not adopted this construction because it is liable to generate false positive signals of recrudescence when the recurrent isolate *r* has MOI*>*1, and exhibits alleles matching the paired baseline isolate *b* at a subset of loci.

### A.2 Modelling allele frequencies

Here, we address the derivation of allele frequencies for both populations *I* and *C*. By convolving Equation (13) over the modelled distribution of population allele frequencies, we show that the locus-wise probability of observations 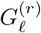 for isolate *r* can be formulated as a multinomial expression of sample allele counts (i.e, allele counts summed over one or more baseline isolates *b* and *i* = 1, …, *n*). The consequences of genotyping error for baseline isolates relative to the recurrent isolate *r*, specified by the probability that alleles called in baseline isolates *b* and *i* = 1, …, *n* match alleles called in the recurrent isolate *r*, will be addressed in Section A.3 in the sub-model of per-isolate allele counts.

#### A.2.1 Allele frequencies for population *I*

Treating isolates 1, …, *n* as a finite sample from a larger population *I*, we derive allele frequencies for population *I* under a Bayesian multinomial-Dirichlet model. We weight each isolate by its MOI, whereby each clone in isolates *i* = 1, …, *n* is weighted equally.

For notational convenience, we introduce an arbitrary ordering on *H*_*𝓁*_, the set if possible alleles at locus *𝓁*. For the *k*^th^ allele *α*_*k*_ in the set *H*_*𝓁*_, we denote by 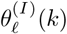 the baseline population frequency, and by

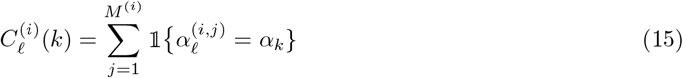

the per-isolate allele count ∈ *{*0, 1, …, *M* ^(*i*)^*}* across clones in isolate 1, …, *n*.

Under the assumed independence of clones within and between the baseline isolates 1, …, *n*, we model sample allele counts aggregated over isolates 1, …, *n* to follow a multinomial distribution

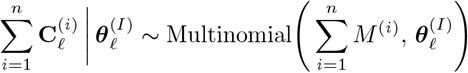

governed by the population allele frequencies 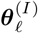.

Taking uniform prior for 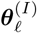 over the |*H*_*𝓁*_| − 1 simplex, Dirichlet |*H*_*𝓁*_|, **1**, the posterior distribution of allele frequencies 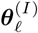 given the per-isolate allele counts 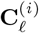 takes the form

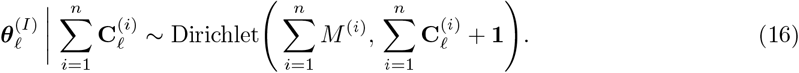

#### A.2.2 Allele frequencies for population *C*

Allele frequencies for population *C* are largely governed by clones in the paired baseline isolate *b*. However, we accommodate undetected clones in isolate *b*, with allelic states for ungenotyped clones imputed using population *I* allele frequencies 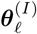. This imputation is the rationale for withholding the paired isolate *b* when deriving population *I* allele frequencies.

Akin to the model of (un)genotyped clones for isolate *r* (Equation (11)), we model each clone in isolate *b* to be detected at locus *𝓁* with probability *ω*. Then the number of clones 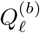 in isolate *b* that are successfully genotyped at locus *𝓁* (that is, contribute to the observation 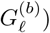 is modelled to follow a truncated binomial distribution, with the lower bound of the support given by the observed cardinality 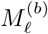 at locus *𝓁*:

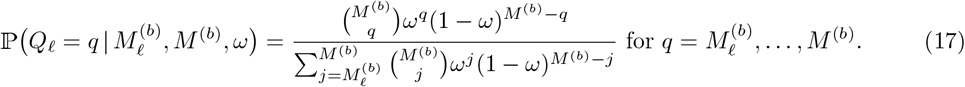

Given 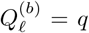, let 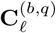 be the vector of allele counts across the *q* clones in isolate *b* that have been successfully genotyped at locus *𝓁*. Then, conditional on 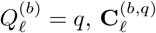 and 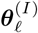, we model

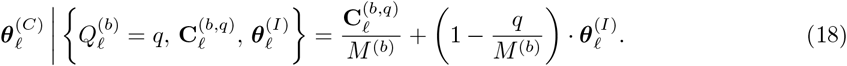

***Remark*** : Assuming perfect detection of clones in the paired baseline isolate *b* (i.e., setting *ω* = 1) can have strong implications when paired recurrences share alleles at all but one locus, with a length difference that cannot plausibly be explained by genotyping error, but likely reflects human error; see Figure 7 for two empirical examples from [17]. Accommodating ungenotyped clones in isolate *b* can prevent a single locus from having a disproportionately large effect on overall classifications.

#### A.2.3 A locus-wise model predicated on allele counts

Using the derived distributions of allele frequencies for population *I*, 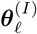 (Equation (16)), and population *C*, 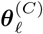 (Equation (18)), we can formulate the locus-wise probability of observations 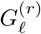 for isolate *r* in terms of sample allele counts.

Substituting Equation (18) into Equation (13) and convolving over the truncated binomial distribution 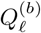 (17) governing the number of clones detected at locus *𝓁* in isolate *b*, we find that:

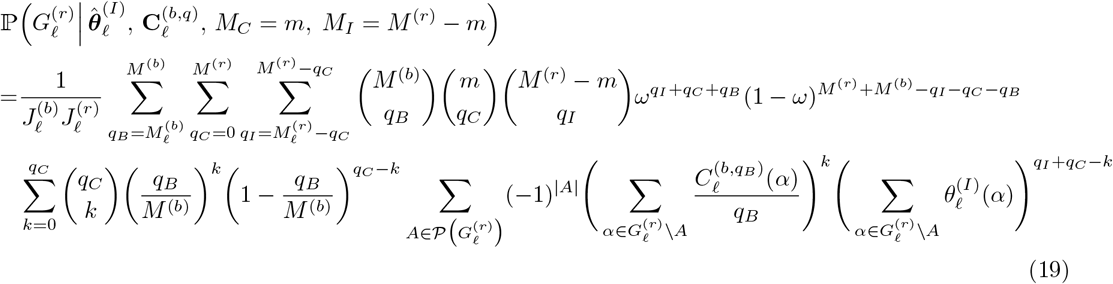

where we set

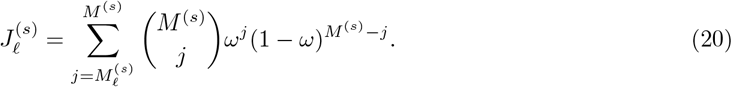

Convolving Equation (19) over the derived distribution of 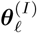 (16) (with integration performed over the |*H*_*𝓁*_| − 1 simplex) yields the locus-wise probability of observations 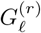 for isolate *r* formulated with respect to the per-isolate allele counts 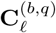 across successfully genotyped clones in isolate *b*, and the sample allele counts 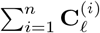:

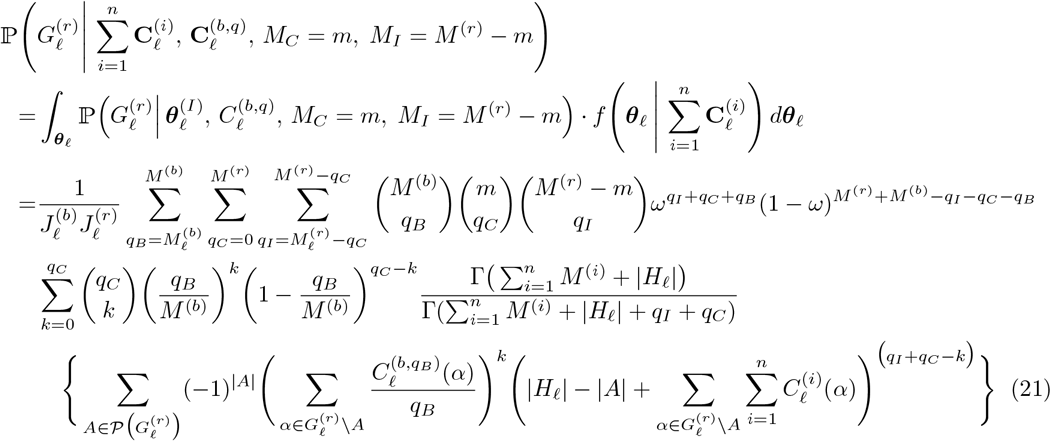

where (*·*)^(*·*)^ denotes the rising factorial. Here, we have used the aggregation property of the Dirichlet distribution.

To write Equation (21) in terms of simple powers of 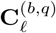 and 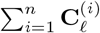, we apply the binomial expansion to yield

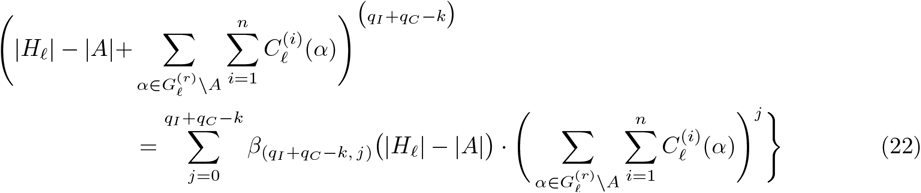

where the coefficients

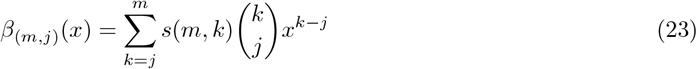

are formulated with respect to unsigned Stirling numbers of the first kind *s*(*m, k*). Substituting Equation (22) into Equation (21) yields a multinomial expression of the per-isolate allele counts **C**^(*b,q*)^ and the sample allele counts 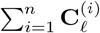, as desired.

## A.3 Moments of allele counts

Using bulk genotypic data, allele counts are not directly observed for multiclonal isolates. To recover the locus-wise likelihood of observations 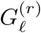 for isolate *r* given it comprises *M*_*C*_ = *m* recrudescent and *M*_*I*_ = *M* ^(*r*)^ − *m* newly-inoculated clones, we must convolve Equation (21) over the underlying isolate-wise distribution of allele counts. Because the locus-wise probability of observations 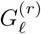 for isolate *r* can be written as a multinomial expression of the per-isolate allele counts **C**^(*b,q*)^ and sample allele counts 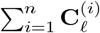 (Equations (21) and (22)), evaluation of the locus-wise likelihood distils down to the computation of *moments* of these allele counts. This is a tractable combinatorial problem which can be solved explicitly under our simplifying assumptions.

Substituting Equation (22) into Equation (21) and convolving over the distribution of per-isolate allele counts for baseline isolates *b* and *i* = 1, …, *n*, we obtain

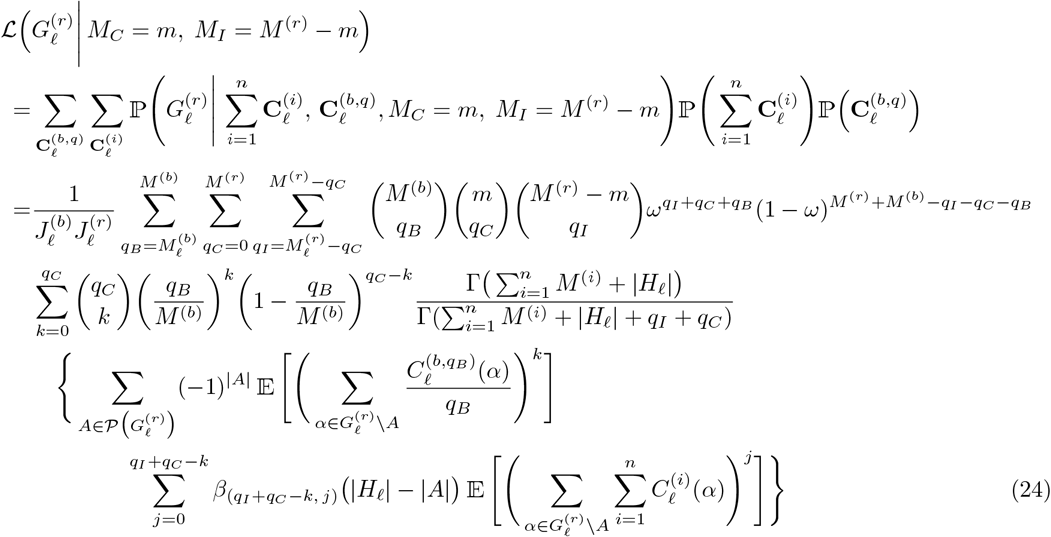

where we have evoked the assumed independence of (genotyped) clones across baseline isolates *b* and 1,.

Evaluating the locus-wise likelihood of observations 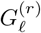 for isolate *r* (24) necessitates the calculation of moments of the collective counts of allelic subsets *A*^*′*^ summed over one or more baseline isolates. We can derive these moments explicitly under three key assumptions:

1. We consider the distribution of per-isolate allele counts 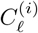 for each isolate *i* = 1, …, *n*, such that all of the *M* ^(*i*)^ clones in isolate *i* contribute to the observation 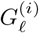 at locus *𝓁*, whereby we assume implicitly that the effects of ungenotyped clones (if any) ‘average out’ across isolates *i* = 1, …, *n* when formulating allele frequencies for population *I*; note that 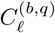 corresponds explicitly to the *Q*_*𝓁*_ = *q* clones within isolate *b* that have been successfully genotyped at locus *𝓁* and therefore contribute to the observation 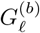.
2. We model clones to be independent within and between baseline isolates.
3. We take a uniform distribution over compatible allelic configurations within each isolate, that is, any allocation of the alleles 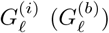 over 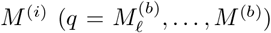 clones, such that at least one clone harbours each allele in the set 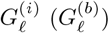, is modelled to be equally likely.

As foreshadowed, we adjust allele counts in baseline isolates for genotyping error relative to the recurrent isolate *r*, in accordance with the non-parametric model *δ*_*𝓁*_(*α, α*^*′*^) which yields the probability that an allele identified as *α* ∈ *H*_*𝓁*_ in a baseline isolate matches allele *α*^*′*^ ∈ *H*_*𝓁*_ in a recurrent isolate *r*.

### A.3.1 Per-isolate allele counts

We begin by characterising per-isolate allele counts 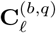 for the *q* clones in isolate *b* that are successfully genotyped at locus *𝓁*. Denote by

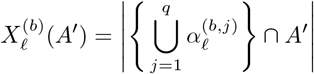

the number of distinct alleles, harboured by the *q* clones in isolate *b* that are successfully genotyped at locus *𝓁*, that lie in a specified set *A*^*′*^ ⊂ *H*_*𝓁*_. Accommodating genotyping error *relative* to the recurrent isolate *r*, each observed allele 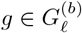 matches an allele within the set 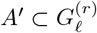 with probability

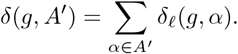

We model genotyping error independently for each allele in the observed set 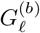. Accordingly, we model 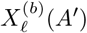 as a sum of 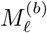 independent but not identically distributed Bernoulli random variables (i.e., a Poisson binomial distribution) with respective success probabilities 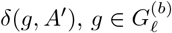.

Define

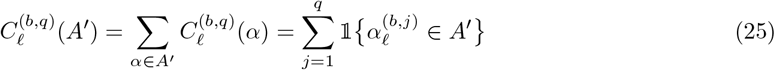

to be the number of clones within isolate *b* that are genotyped (correctly or incorrectly) at locus *𝓁* and harbour an allele in the set *A*^*′*^ at locus *𝓁*. We can recover the distribution of 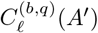 using a simple combinatorial argument as follows.

The number of allelic configurations 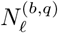 of *q* clones compatible with the observation 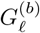 at locus *𝓁* is equivalent to the number of ordered partitions of *q* objects into 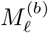 non-empty subsets. We can write this in the form

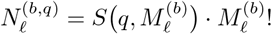

where *S*(*n, k*) denotes the Stirling number of the second kind.

Given 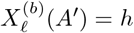, allelic configurations of *q* clones such that 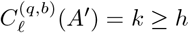 clones harbour alleles in the set *A*^*′*^ can be generated by:

- Separating the *q* clones into two groups of size *k* and *q* − *k* respectively, which can be performed in 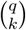 ways;
- For the group of size *k* (corresponding to clones with alleles in the set *A*^*′*^), constructing ordered partitions into *h* non-empty subsets, which can be performed in *S*(*k, h*) *· h*! ways;
- For the group of size (*q* − *k*) (corresponding to clones with alleles outside the set *A*^*′*^), constructing ordered partitions into 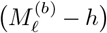 non-empty subsets, which can be performed in 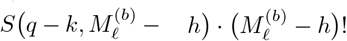 ways.

Therefore, adjusting for genotyping error relative to the recurrent isolate *r*, the number of allelic configurations of *q* clones, such that the unique set of observed alleles at locus *𝓁* is 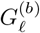 *and* precisely 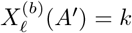 clones harbour alleles within the set *A*^*′*^, takes the form

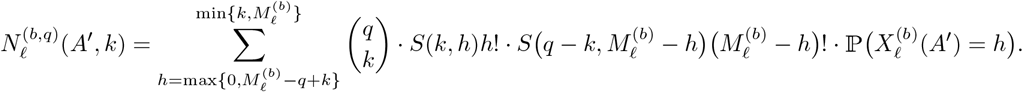

Efficient computation of the probability mass function 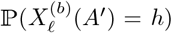 of the Poisson binomial distribution can be performed using the DFT method of Hong [43], with an implementation available in the R package poisbinom [38]. The consequences of genotyping error, relative to the recurrent isolate *r*, are embedded implicitly within the term 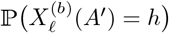.

We thus obtain the probability mass function for the number of clones 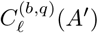 (of the *q* clones that are genotyped at locus *𝓁* in isolate *b*) with alleles lying in the set *A*^*′*^ at locus *𝓁*:

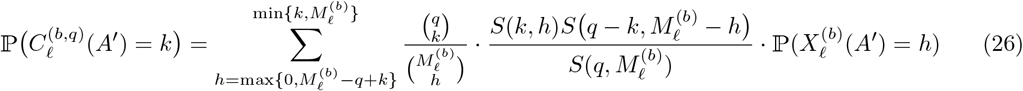

for *k* ∈ *{*0, 1, …, *q}*. We can use the PMF to (26) to compute moments of *C*^(*b,q*)^(*A*^*′*^) for relevant allelic subsets *A*^*′*^ to be substituted into Equation (24).

#### Sample allele counts

The likelihood (24) additionally requires computation of the moments of sample counts of allelic sets *A*^*′*^:

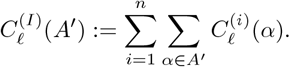

Here, we exploit the fact that the order of these moments (up to *M* ^(*r*)^) is likely to be smaller than the size of the baseline sample *n*. With minor modifications in notation, we can use the PMF (26) for per-isolate allele counts to directly compute the *y*^th^ cumulant 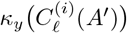 for the number of clones 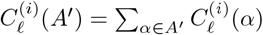 within each isolate *i* = 1, …, *n* that harbour alleles within the set *A*^*′*^. Due to the independence of clones across baseline isolates, it follows that

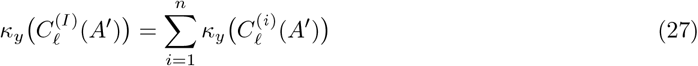

To recover the *m*^th^ moment of 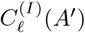, we exploit the mapping between moments and cumulants:

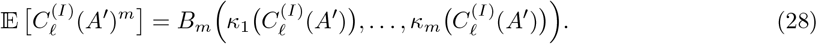

where *B*_*m*_(*·*) denotes the *m*^*th*^ complete exponential Bell polynomial. Equation (28) can then be substituted into the likelihood (24) for relevant allelic subsets *A*^*′*^.

## B Validation against simulated data

To validate our classification model *PfRecur*, we simulate recurrent isolates as mixtures of newly-inoculated and recrudescent clones. A summary of simulation parameters is provided in Table 3.

**Table 3.** Summary of simulation parameters.

| Parameter | Interpretation | Value |
| --- | --- | --- |
| $L$ | Number of independent (unlinked) and neutral markers | 7 |
| $\mathbf{h}$ | Vector of marker cardinalities (number of possible alleles $\vert H_\ell\vert $ at each marker $\ell = 1, \dots, L$) | (30, 30, 20, 20, 20, 10) |
| $\varepsilon$ | Probability of genotyping error per allele call in each baseline isolate | 0.05 |
| $M_{\max}$ | Maximum MOI (number of not necessarily distinct clones) within each isolate | 9 |
| $\mu$ | Mean of the geometric distribution (with non-zero state space) governing the isolate MOI (truncated at $M_{\max}$) | 3 |
| $p_{\text{sib}}$ | Parameter governing within-isolate sibling structure (represented by non-crossing partitions); zero corresponds to the absence of sibling structure, i.e., independent clones | 0.1 |
| $\omega$ | Per-clone marker-wise probability of detection | 0.9 |
| $n$ | Number of baseline isolates | 25 |

**Table 4.** Efficacy estimates without PCR correction (as reported in Table 4 of [17]) vs based on posterior probabilities of recrudescence under the CDC model vs metric M1 under *PfRecur* (with default parameters *ω* = 0.9, *ε* = 0.05), adjusted for loss to follow-up under a Kaplan-Meier model [8, 17].

|  | Study site | Uncorrected | CDC model | *PfRecur* |
| --- | --- | --- | --- | --- |
| **AL** (*day 28*) | Zaire | 77.1 (69–86) | 88.9 (82.8–95.3) | 90.4 (84.5–96.4) |
|  | Lunda Sul | 87.3 (81–94) | 94.4 (89.9–99.0) | 96.3 (92.6–100) |
| **ASAQ** (*day 28*) | Zaire | 82.9 (76–91) | 93.9 (89.1–98.9) | 94.4 (89.8–99.2) |
|  | Lunda Sul | 100 | 100 | 100 |
| **ASPY** (*day 42*) | Benguela | 87.2 (81–94) | 99.7 (99.2–100) | 100 (100–100) |
| **DP** (*day 42*) | Benguela | 94.2 (90–99) | 98.3 (95.8–100) | 99.0 (97.1–100) |

We generate baseline samples comprising *n* independent isolates, accompanied by paired recurrent isolates. For each isolate, we simulate genotypic data across *L* markers, indexed *𝓁* = 1, …, *L*.

Each marker *𝓁* is uniquely specified by a cardinality *h*_*𝓁*_. The set of possible alleles at marker *𝓁* is denoted *H*_*𝓁*_ = *{*1, …, *h*_*𝓁*_*}*, with the integer label interpreted as a length (in arbitrary units).

We accommodate structured genotyping error at each marker *𝓁*. We adopt a normalised geometric model, based on the absolute length difference between an observed and underlying allele [8], with success parameter (1−*ε*). We normalise this model such that each allele is erroneously genotyped with probability *ε*, that is,

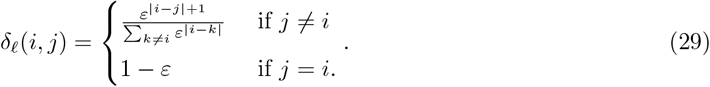

We then generate a random permutation *σ* of the set *H*_*𝓁*_. The population frequency of the *σ*(*i*)^th^ allele at marker *𝓁* is taken to be

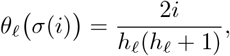

yielding the effective cardinality [44]

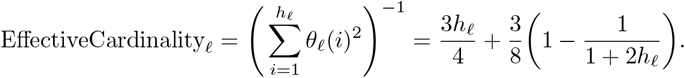

This is motivated by empirical data, which appear to yield an approximately-linear trend in ordered (expected) sample allele frequencies.

The MOI of each isolate (baseline or recurrent) is sampled from the geometric distribution with mean *µ* and non-zero state space, truncated at *M*_max_, that is,

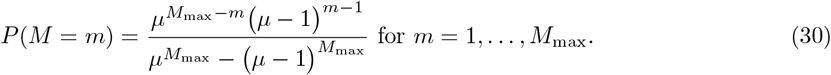

We accommodate the presence of siblings within simulated isolates. To generate a baseline set of clones for isolate *i* with MOI *M* ^(*i*)^, we first simulate a non-crossing partition of the set *{*1, …, *M* ^(*i*)^*}*, governed by the partition parameter *p*_sib_: given

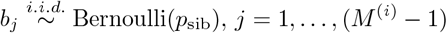

clones *j* and (*j* + 1) are assigned to the same block if and only if *b*_*j*_ = 0. For each block of siblings *B*, we then generate a pair of independent parents. The allelic state 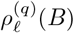 of parent *q* ∈ *{*1, 2*}* of block *B* at locus *𝓁* is sampled from a categorical distribution governed by the population allele frequencies

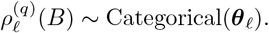

We then recover each clone in block *B* as a mosaic of these independent parents, retaining the assumption of unlinked markers: for each clone *j* ∈ *B*, the allelic state at marker *𝓁*

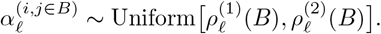

For each clone *j* = 1, …, *M* ^(*i*)^, we simulate a Bernoulli random variable

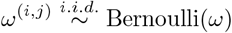

governing detection due to genotyping at locus *𝓁*. We then aggregate the set of detected alleles present at locus *𝓁*

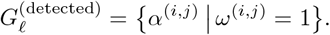

For each detected allele 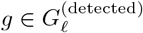, we sample an observed allele from the categorical distribution

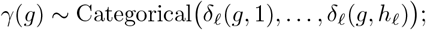

classification and population allele frequency estimation is performed based on the observed set of genotypes 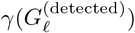 for the baseline isolate.

For the *i*^th^ baseline isolate of MOI *M* ^(*i*)^, we generate a set of paired recurrences as follows. For each 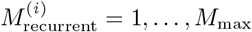 and 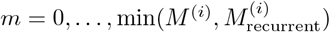, we simulate a mixture of *m* recrudescent and 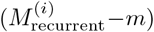 newly-inoculated clones. The *m* recrudescent clones are sampled uniformly at random without replacement from the paired baseline isolate; the 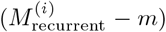 newly-inoculated clones are simulated under the same framework as a baseline isolate. We account for the incomplete detection of clones in the recurrent isolate (with the per-clone marker-wise probability of detection *ω*), but do not account for genotyping error for the recurrent isolate.

Paired simulated recurrences are classified using our novel model *PfRecur*, with a uniform prior over the number of newly-inoculated vs recrudescent clones (i.e., with *β* = 1). Classification is based on the maximum observed cardinality across simulated markers, rather than the true MOI of each isolate. We perform classification in an idealised setting with the user-specified parameters *ω* = 0.9 and *ε* = 0.05 set to the values under which data were simulated.

Results in the main text are aggregated across 40 simulation runs (i.e., 40 simulated sets of baseline samples), amounting to 28773 simulated recurrent isolates.

## C Empirical data: a re-analysis of Dimbu *et al*. [17]

### C.1 Within-host diversity

To ascertain the contribution of each microsatellite marker to the MOI of a given isolate, we derive a within-host diversity metric. For an isolate with MOI *M* ^(*i*)^ and cardinality 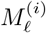 at marker *𝓁*, we define the within-host diversity of marker *𝓁* to be the expected value of the complement of Nei’s gene identity metric (or equivalently, the probability of differing alleles at marker *𝓁* when a pair of clones is sampled with replacement from the isolate), taking a uniform distribution over all possible allelic configurations of *M* ^(*i*)^ clones that harbour precisely 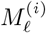 distinct alleles at locus *𝓁*. Following the reasoning in Appendix A.3.1, this takes the form

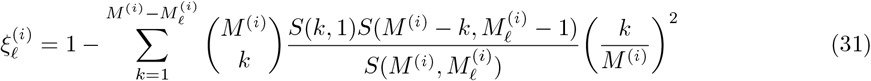

where *S*(*·, ·*) denotes a Stirling number of the second kind.

### C.2 Sensitivity of *PfRecur* to user-specified parameters

Here, we consider the sensitivity of our classification model *PfRecur* to the user-specified parameters *ε* (governing the genotyping error probability under the normalised geometric model (7), applied to each allele called in a baseline isolate relative to the recurrent isolate of interest) and *ω* (governing the marker-wise probability of detection for each clone in the paired baseline and recurrent isolates of interest). A complete sensitivity analysis is available in an accompanying GitHub repository (https://github.com/somyamehra/PfTreatmentFailure); below, we highlight isolates for which classification is sensitive to user-specified parameters.

#### C.2.1 Classification of recurrences in Dimbu *et al*. [17]

The imperfect detection of clones in the paired baseline and recurrent isolate proves a key consideration for a subset of recurrences in Dimbu *et al*. [17] using *PfRecur*. The posterior probability of at least one recrudescent clone (metric M1) is inflated as ungenotyped clones are increasingly accommodated (i.e., the per-clone marker-wise probability of detection *ω* is decreased, Figure 7A). However, accommodating undetected clones in the paired baseline isolate protects against plausible human error, such as the incorrect entry of a single allele in a putative recrudescence (Figure 7B). Inferred mixtures of newly-inoculated vs recrudescent clones, quantified through the posterior proportion of recrudescent clones (metric M2), are also sensitive to per-clone marker-wise probability of detection *ω* in several cases (Figure 7C). A dependence on the genotyping error probability *ε*, under the normalised geometric model (7), is also apparent for a subset of isolates (Figure 8); however, our results are largely insensitive to genotyping error probability within the plausible range 0.01 ≤ *ε* ≤ 0.1. By default, we set *ε* = 0.05 and *ω* = 0.9.

**Figure 7.**
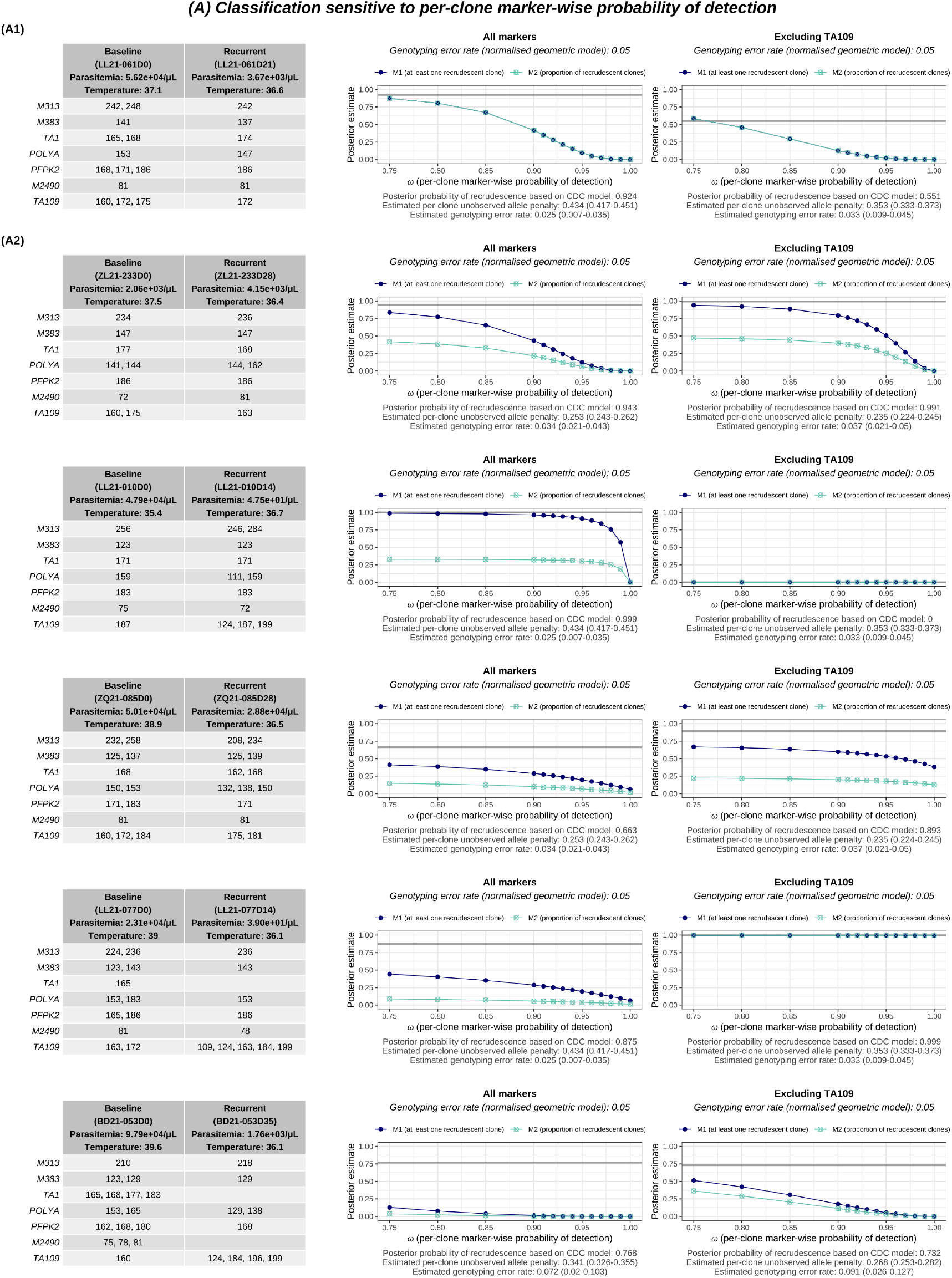
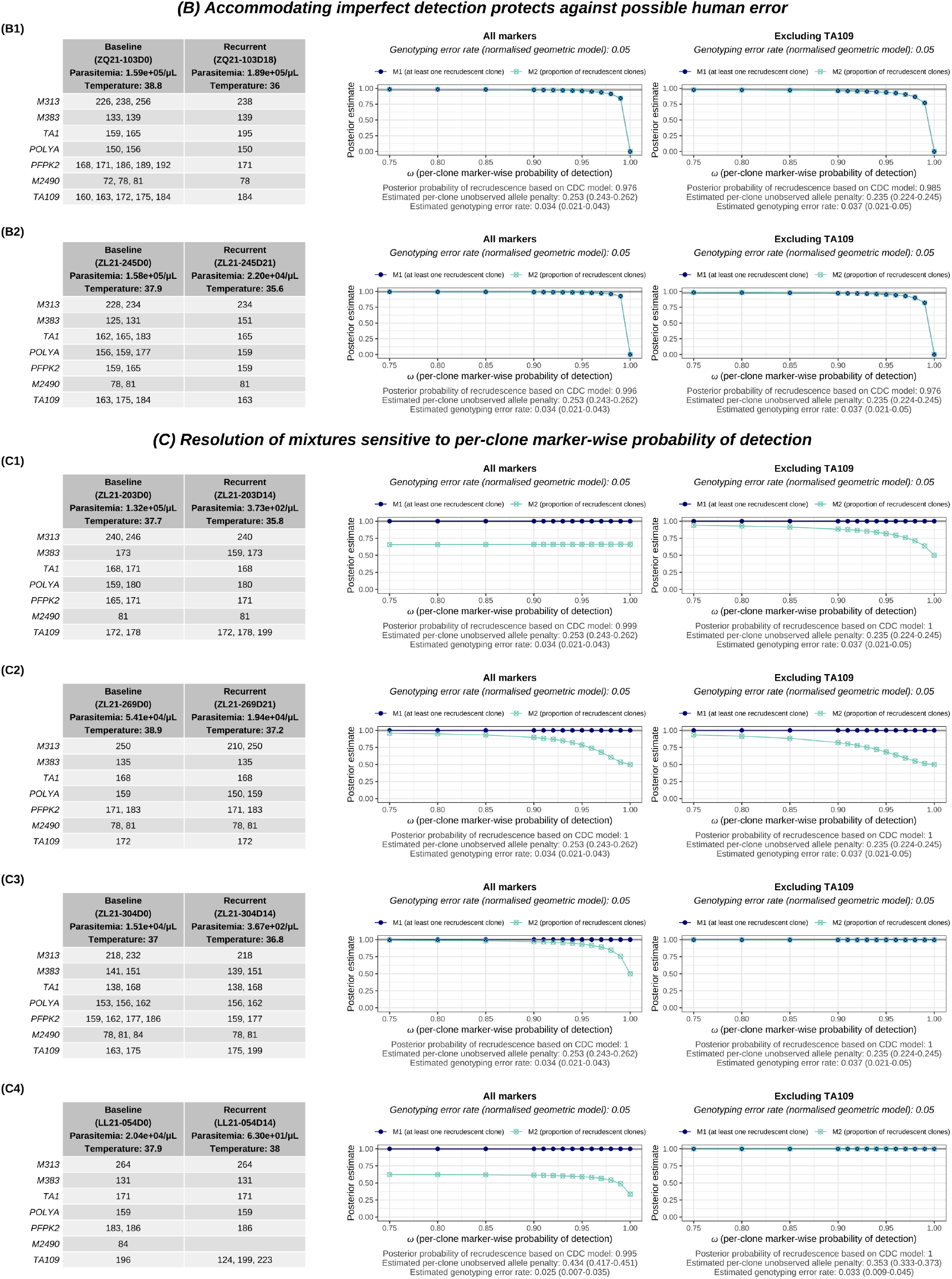
Classifications under *PfRecur* sensitive to the per-clone marker-wise probability of detection ω in the paired baseline/recurrent isolates.

**Figure 8.**
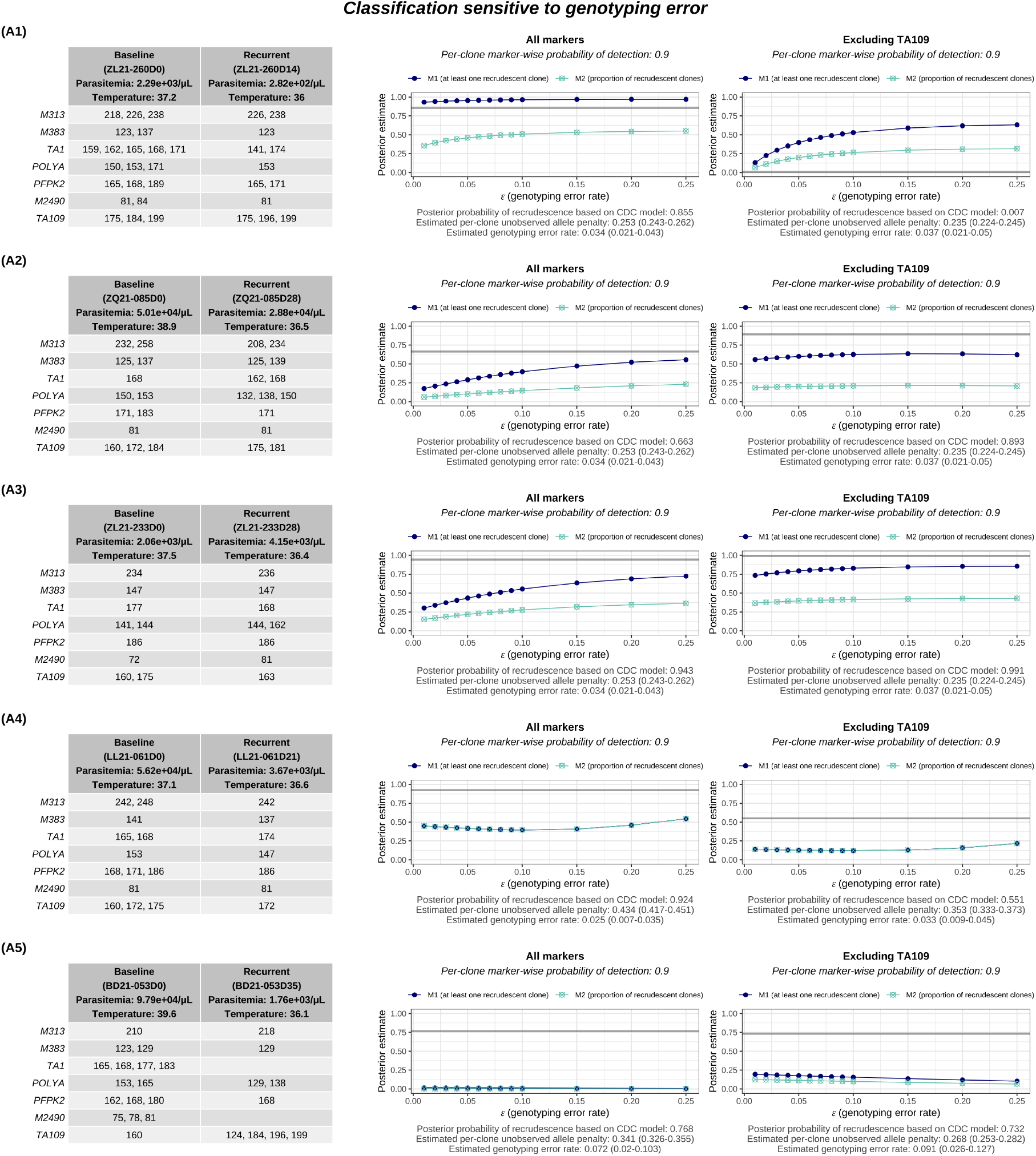
Classifications under *PfRecur* sensitive to the genotyping error probability *ε* (applied to each allele called in a baseline isolate relative to the recurrent isolate of interest) under the normalised geometric model (7).

#### C.2.2 False positive recrudescence rates

Relaxing the per-clone marker-wise probability of detection *ω* can amplify the posterior probability of recrudescence under *PfRecur*. However, false positive recrudescence rates across 500 permuted artificial ‘not-recrudescence’ datasets (obtained by generating derangements of baseline study participant labels within each study site in [17]) remain consistently low for *ε* = 0.05 and *ω* ∈ [0.75, 1]. Empirical complementary CDFs for metric the posterior probability of recrudescence (under the CDC model) vs metric M1 (under *PfRecur*) using which false positive rates are computed are shown in Figure 10.

**Figure 9.**
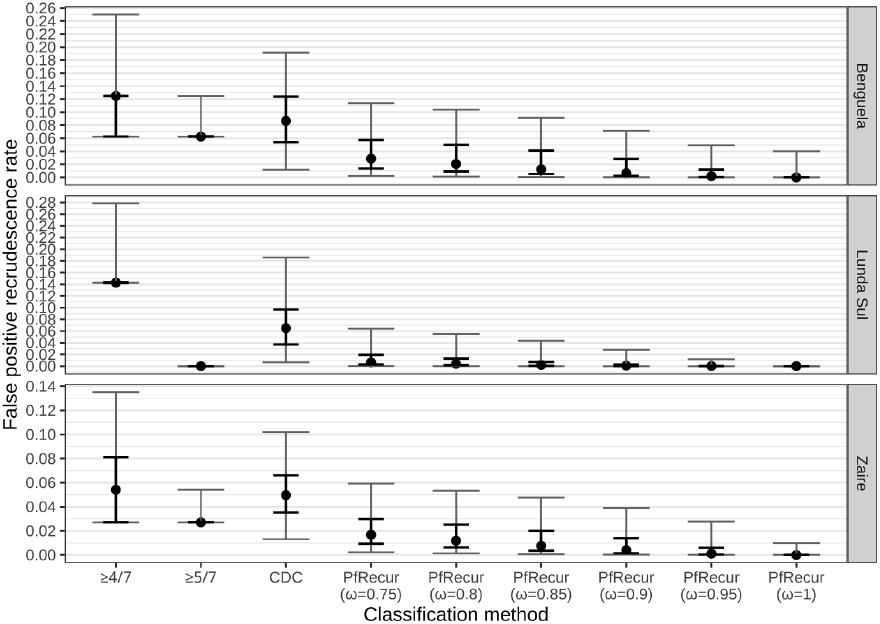
False positive recrudescence rates across 500 permuted artificial ‘not-recrudescence’ datasets under *PfRecur* (metric M1) with genotyping error probability *ε* = 0.05 and per-clone marker-wise detection probability *ω* ∈ [0.75, 1] vs the CDC model vs a match-counting approach. Points show medians; bold error bars show the interquartile range; grey error bars show 95% confidence intervals.

**Figure 10.**
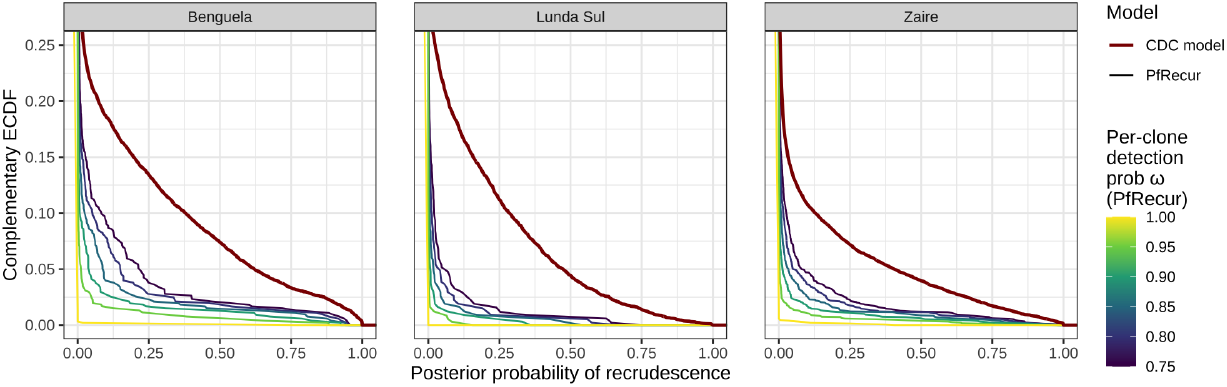
Complementary empirical CDFs for the posterior probability of recrudescence (CDC model) vs metric M1 (*PfRecur*) aggregated over 500 permuted artificial ‘not-recrudescence’ datasets.

### C.3 Comparison against CDC model

We find that the CDC model [8] typically yields elevated posterior probabilities of recrudescence for multiclonal isolates with matching alleles at a subset of markers. Recurrences with differing classifications under *PfRecur* vs the CDC model [8] are summarised in Figure 11.

**Figure 11.**
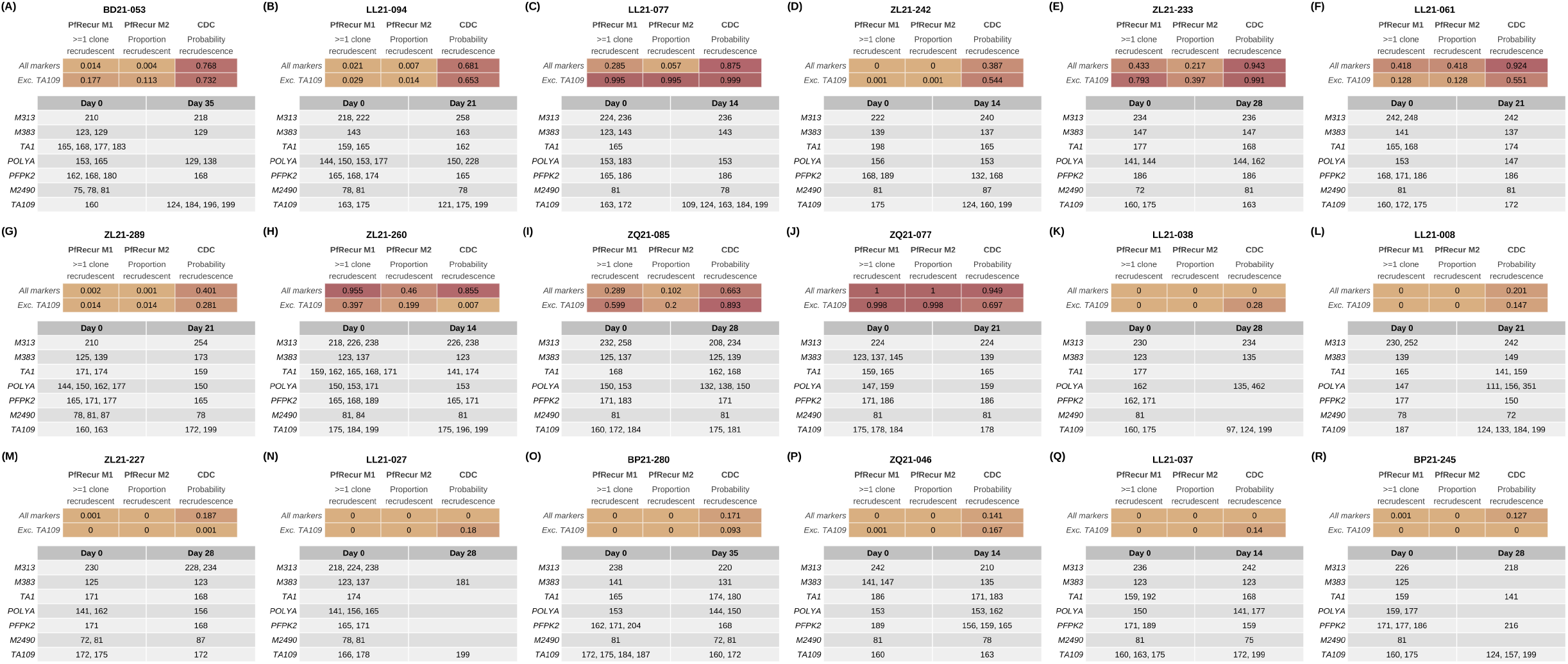
Differing classifications under *PfRecur* (metrics M1 and M2) vs the CDC model [8]. For *PfRecur*, we take a genotyping error probability of *ε* = 0.05 under the normalised geometric model (29) (applied to each allele called in each baseline isolate), and assume that the per-clone marker-wise probability of detection (applied to clones in the paired baseline/recurrent isolates) is *ω* = 0.9.

#### C.3.1 Efficacy estimates

To generate efficacy estimates adjusted for classification uncertainty and right censoring, we perform the Kaplan-Meier correction used in Dimbu *et al*. [17]^3^: *for each study arm, we perform 10,000 iterations of bootstrap resampling whereby we sample a binary reinfection or recrudescence state for each patient under a Bernoulli distribution with success parameter given by the posterior probability of recrudescence (or metric M1 under PfRecur*); and then compute Kaplan-Meier estimates (including the lower 2.5% and upper 97.5% confidence interval under the peto method implemented in the R function survival::survfit [45]) for these binary infection states. Survival estimates for the last day of follow-up (28 days for AL and ASAQ; 42 days for DP and ASPY) are then averaged across these 10,000 iterations to yield an adjusted efficacy estimate and an accompanying 95% confidence interval. Efficacy estimates under *PfRecur* vs the CDC model (re-run on our system) are summarised in Table 4. Efficacy estimates generated using the CDC model in Table 4 differ from those reported in Dimbu *et al*. [17] due to the stochastic nature of the MCMC algorithm, and because early treatment failures (1 patient treated with AL, and 3 patients treated with ASAQ in Zaire) have not been taken into account in our re-analysis.

## D Comparison against CDC model structure [8]

An extended comparison of the *PfRecur* model structure against the CDC model [8] is provided below. In brief, the CDC model [8] explicitly estimates allelic configurations using a Gibbs sampler; averages the likelihood of recrudescence across pairs of clones in a baseline and recurrent isolate; and concurrently generates estimates of parametric (geometric) genotyping error probabilities and a per-clone unobserved allele “penalty”. In contrast, in *PfRecur* we average analytically over compatible allelic configurations in each isolate to obtain an analytically tractable posterior; model recurrent isolates as mixtures of newly-inoculated vs recrudescent clones; and treat both genotyping error and the per-clone marker-wise probability of detection for paired baseline/recurrent isolates as user-specified parameters.

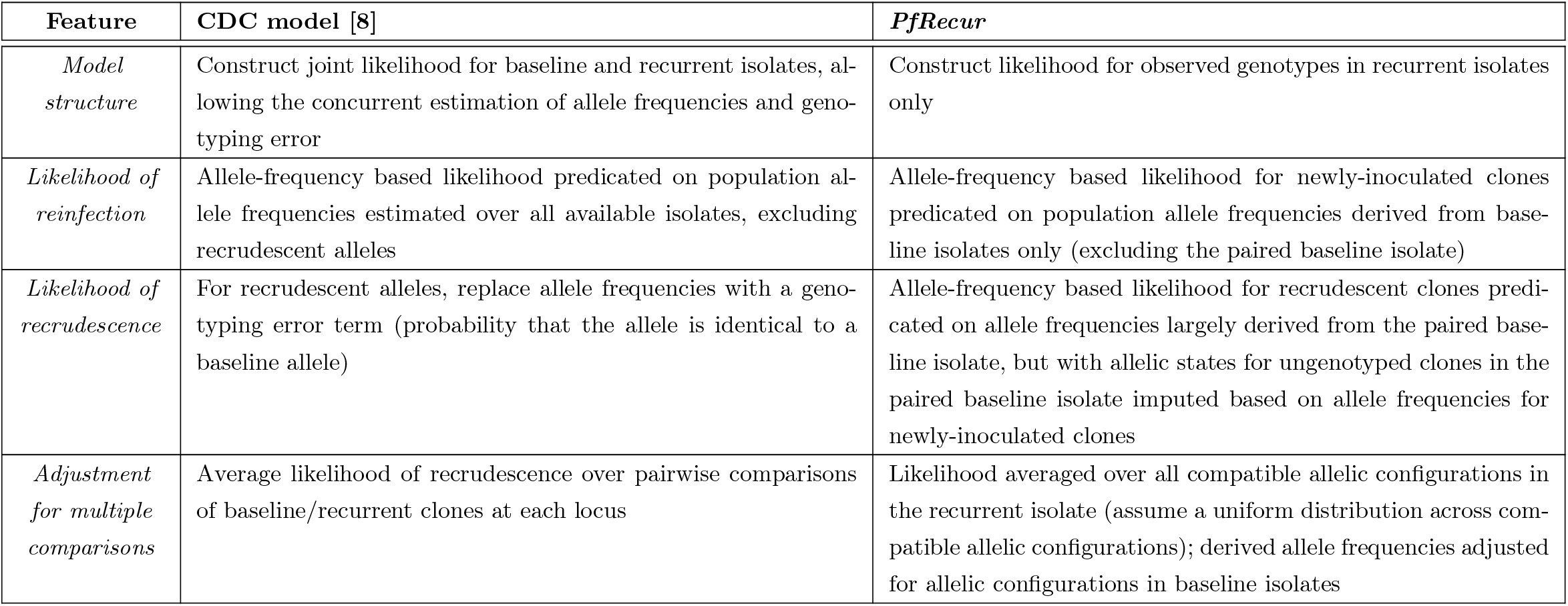
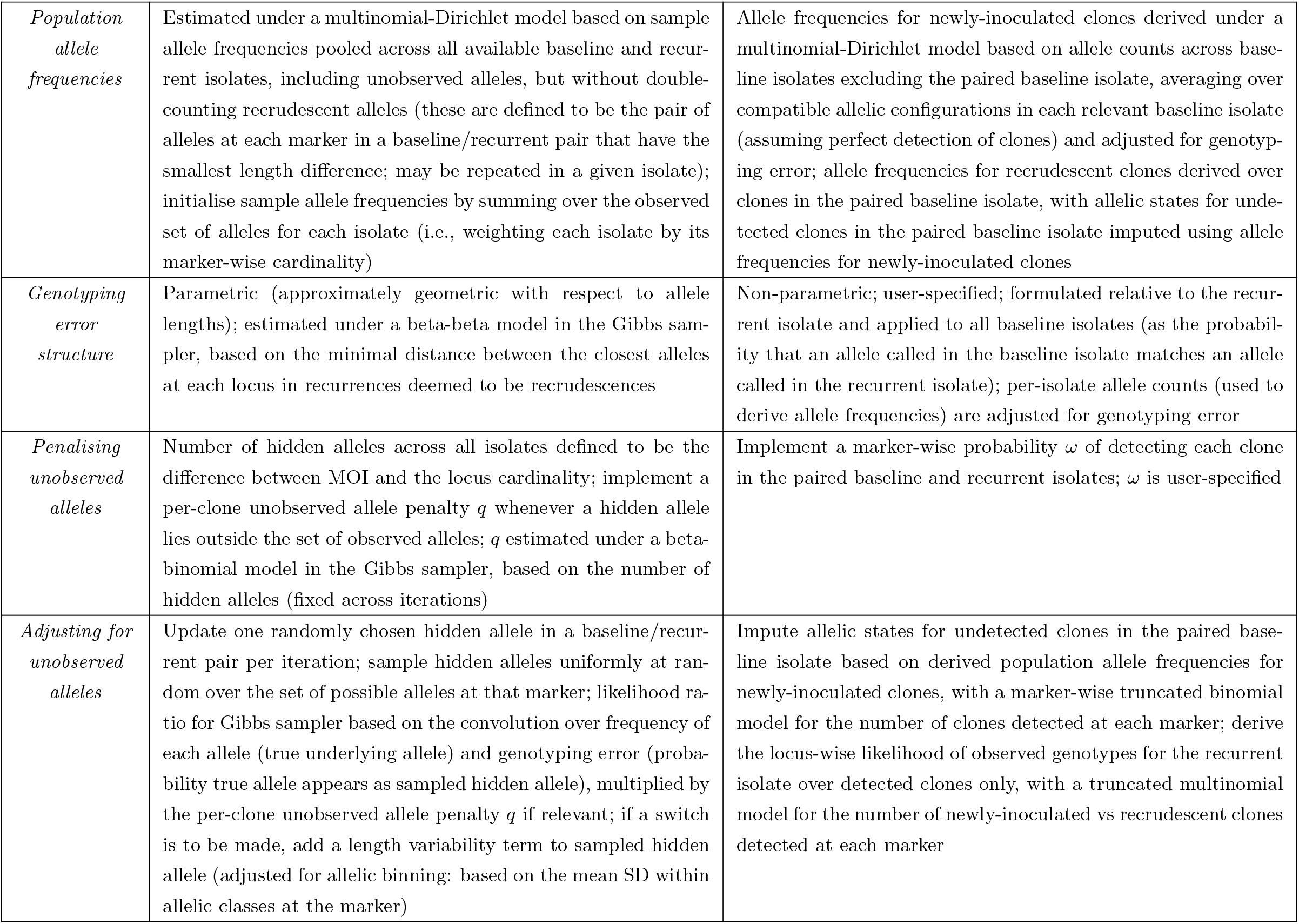

## Footnotes

1 Measures of allelic abundance are not readily available for length polymorphic markers that are genotyped using electrophoresis and currently used to perform molecular correction. However, amplicon sequencing, which is recognised by the WHO as a potential future standard for molecular correction, generates read count data on multi-allelic markers (microhaplotypes), and read count data can be used to estimate the relative abundance of within-isolate alleles.

2 Current WHO guidelines, tailored to a 3 marker panel, stipulate a strict 3/3 match-counting rule for primary analysis [9]. We do not consider the strict 7/7 match-counting rule for the 7 neutral microsatellite panel used in [17], given the microsatellite marker TA109 appears to be problematic. There are also several recurrences in [17] (namely, ZL21-292, ZQ21-103 and ZL21-245) where mismatch at a single marker (with a length difference plausibly attributable to either genotyping or human error) supports the use of a relaxed match-counting rule.

3 Implemented in https://github.com/MateuszPlucinski/AngolaTES2021 [40]

## Notes

### Competing Interest Statement

The authors have declared no competing interest.

### Author Declarations

This study uses open access data that has previously been published by Dimbu et al (2024) (DOI:10.1128/aac.01525-23). Parasite densities and clinical metadata have been retrieved from Supplemental Table S4 of Dimbu et al (2024), while genotypic data have been retrieved from an accompanying GitHub repository (https://github.com/MateuszPlucinski/AngolaTES2021).

